# Network Meta-Analysis Comparing the Short and Long-Term Outcomes of Alternative Access for Transcatheter Aortic Valve Replacement

**DOI:** 10.1101/2021.09.06.21263150

**Authors:** Sagar Ranka, Shubham Lahan, Adnan K. Chhatriwalla, Keith B. Allen, Sadhika Verma, Pedro Villablanca

## Abstract

**Objectives:** This study aimed to compare short- and long-term outcomes following various alternative access routes for transcatheter aortic valve replacement (TAVR).

**Methods:** Thirty-four studies with a pooled sample size of 30,986 records were selected by searching PubMed and Cochrane library databases from inception through 11^th^ June 2021 for patients undergoing TAVR via 1 of 6 different access sites: Transfemoral (TF), Transaortic (TAO), Transapical (TA), Transcarotid (TC), Transaxillary/Subclavian (TSA), and Transcaval (TCV). Data extracted from these studies were used to conduct a frequentist network meta-analysis with a random-effects model using TF access as a reference group.

**Results:** Compared with TF, both TAO [RR 1.91, 95% CI (1.46–2.50)] and TA access [RR 2.12, 95%CI (1.84–2.46)] were associated with an increased risk of 30-day mortality. No significant difference was observed for stroke, myocardial infarction, major bleeding, conversion to open surgery, and major adverse cardiovascular or cerebrovascular events in the short-term (≤ 30 days). Major vascular complications were lower in TA [RR 0.43, (95% CI, 0.28-0.67)] and TC [RR 0.51, 95% CI (0.35-0.73)] access compared to TF. The 1-year mortality was higher in the TAO [RR of 1.35, (95% CI, 1.01–1.81)] and TA [RR 1.44, (95% CI, 1.14–1.81)] groups.

**Conclusion:** Non-thoracic alternative access site utilization for TAVR implantation (TC, TSA and TCV) is associated with similar outcomes to conventional TF access. Thoracic TAVR access (TAO and TA) is associated with increased short and long-term mortality.

## Introduction

Transcatheter aortic valve replacement (TAVR) is increasingly performed as a favored treatment modality over surgical aortic valve replacement among patients of all risk profiles with severe aortic stenosis.^1-4^ The transfemoral (TF) access has been most widely used for TAVR valve deployment and is considered as gold standard for the procedure.^1,5-7^ However, in about 10%–15% of patients, TF access is not feasible owing to unfavorable iliofemoral arterial anatomy, necessitating a need for an alternative approach.^8^

The risk profile of patients needing alternate access TAVR is typically higher compared to TF TAVR patients, however, the safety and efficacy of alternative access TAVR as compared to ‘like’ TF TAVR patients remains controversial. The selection of alternate access is often driven by the expertise and make-up of the heart team with options for percutaneous vs surgical access, need for general anesthesia, and potential impact on the cerebral, respiratory and renal systems all playing a role in the selection. Current evidence for alternative access is largely derived from observational studies comparing these alternative access strategies in a ‘pairwise’ fashion.^9-12^ No randomized trial directly compares different approaches and these approaches are more or less subject to a learning curve.^13^ This limits the scope of clinical decision making, creating equipoise, especially when multiple access site options are available. There is an unmet need for a comprehensive analysis evaluating multiple alternative access options simultaneously. Therefore, we conducted this network meta-analysis to evaluate the short- and long-term outcomes of various access approaches for TAVR.

## Methods

We conducted this network meta-analysis in accordance with the preferred reporting items for systematic review and meta-analyses (PRISMA) guidelines and the American Heart Association (AHA) scientific report on methodological standards for systematic reviews and meta-analyses.^14,15^ A PRISMA checklist has been provided in the ***eTable-1, Data Supplement***.

### Data Availability Statement

The authors declare that all the data are available in the article and in its Data Supplement files.

### Data Sources and Searches

Given the public availability of data and the de-identified nature of patient information in respective studies included in the current network meta-analysis, this study was exempt from approval by the institutional review board. Two investigators (S.L. and S.R.) performed the literature search using the PubMed/Medline and Cochrane library databases through 11^th^ April 2021 to identify and retrieve all the studies comparing various approaches for TAVR by using the following search terms: “transcatheter aortic valve replacement”, “transfemoral”, “transaortic”, “transaxillary”, “subclavian”, “mortality”, “transcarotid”, “clinical outcomes”, and “access site.” The same authors independently reviewed 10,098 citations by their titles, of which 5,796 articles were identified as duplicates and were thus discarded. The remaining 4,302 articles were screened by their titles and abstracts, of which 4,157 were excluded, leaving a total of 145 full-text articles to be assessed for eligibility. A total of 111 articles were identified as systematic reviews and meta-analyses, editorial comments, case-reports, duplicate studies, or review articles, and were therefore excluded. One study, by Kirker et al, compared transcarotid TAVR with transaortic/transapical access^16^ but was excluded owing to combined reporting of transaortic and transapical outcomes under “transthoracic” cohort. Finally, 34 studies satisfied our inclusion criteria and were quantitatively evaluated (**Figure 1**). Any conflicts regarding the study selection were resolved by a mutual consensus.

**Figure 1.**
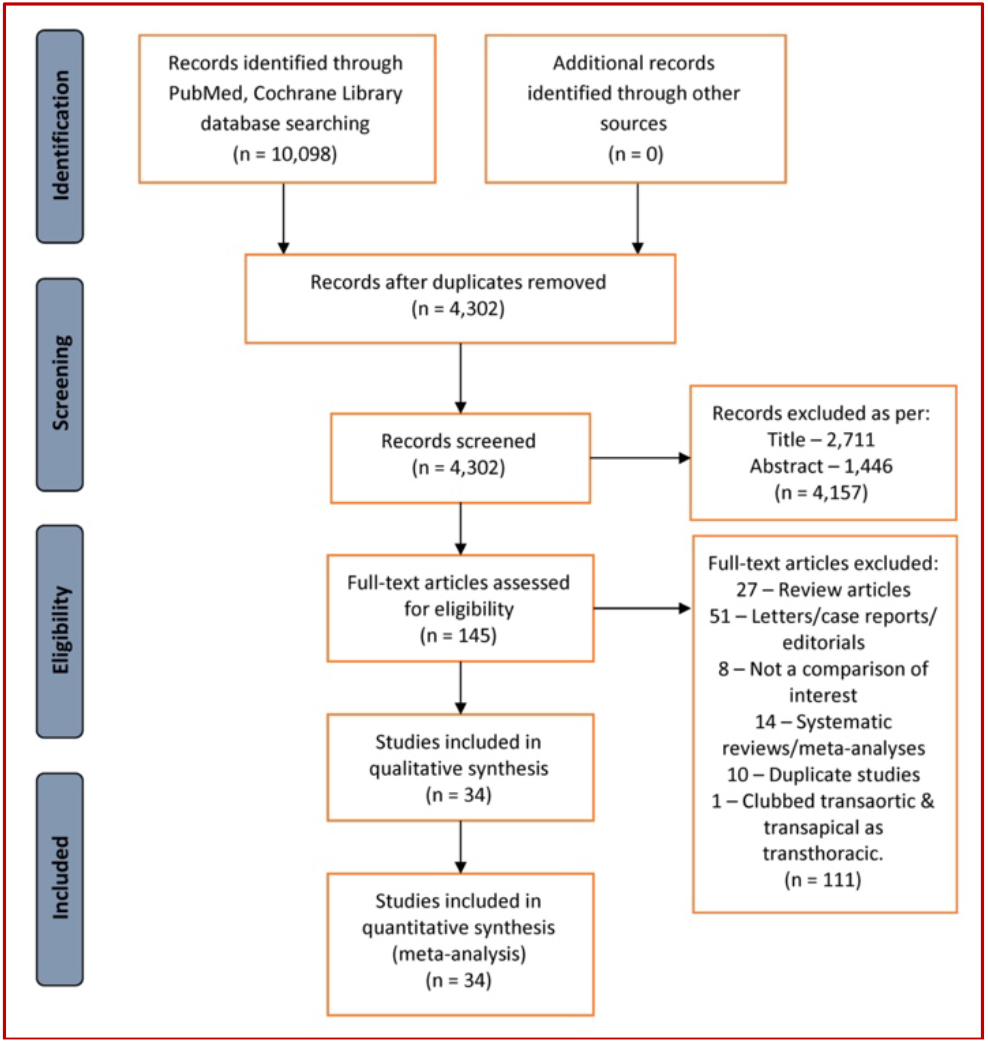
PRISMA diagram illustrating the study selection process.

### Study Selection

We aimed to include all published English language articles (randomized controlled trails, cohort, and observational studies) comparing various approaches of TAVR with at least a 30-day clinical follow-up. A total of 6 TAVR approaches were included: TF, subclavian/axillary (TSA), transcaval (TCV), transcarotid (TC), transapical (TA), and transaortic (TAO).

### Data Abstraction and Quality Assessment

Data on clinical outcomes and study were extracted independently by two authors (S.L. & S.R.) using a standardized data extraction form with an intention-to-treat principle. Any disagreements pertaining to the information were resolved by mutual consensus. We used Methodological Index for Non-Randomized Studies (MINORS) criteria for performing quality assessment.^17^ The criteria consist of 12 methodological parameters for evaluating study quality; with the ideal score being 24 for comparative studies. The overall scores of studies included in this network meta-analysis ranged from 16–22. The details of quality assessment are provided in **eTable-4, Data Supplement**.

### Outcomes

Our pre-defined primary outcomes were short- (30-days) and long-term (≥1-year) all-cause mortality following TAVR. The secondary outcomes included short-term risk of conversion to open cardiac surgery, implantation of a new pacemaker, major vascular complication, stroke, myocardial infarction, major bleeding, and major adverse cardiovascular or cerebrovascular events (MACCE). The outcomes were defined as they were in respective studies included in this network meta-analysis.

### Statistical Analysis

We conducted this frequentist network meta-analysis using a random-effects model. The outcomes were reported as risk ratios (RR) along with their corresponding 95% confidence intervals (CI). We used the P-score metric, which is based entirely on the point estimates and standard errors of the frequentist network meta-analysis estimates for comparing the hierarchy of effectiveness and safety of various treatments. P-score quantitatively ascertains that a particular intervention is better than others averaged over all competing interventions. The values range from 0-1, i.e., the higher the value, the higher the effectiveness of a treatment.^18^ The consistency between the direct and indirect estimates was assessed by using the node-splitting technique (**eTable-5 & eFigure-1, Data Supplement**), and heterogeneity was quantified by tau-square (τ^2^) and I^2^ statistic (**eTable-6, Data Supplement**). Pairwise meta-analyses using DerSimonian and Laird random-effects model reporting direct estimates were also performed (**eTable-7, Data Supplement**). We used Wan method to convert values reported as median (interquartile range) into mean ± standard deviation.^19^ We used *meta* and *netmeta* packages for conducting our network meta-analysis.^20,21^ All statistical analyses were conducted in R (v 4.0.2).

## Results

### Study Search and Characteristics

We identified a total of 34 studies comprising a pooled population of 32,689 patients.^6,7,22-53^ We compared outcomes across 6 different access sites used for TAVR: TF, TSA, TCV, TC, TAO, and TA. **Table 1** details the baseline characteristics of studies and patients belonging to individual studies. The mean age was similar across the groups. Only 22.1% patients who underwent transfemoral TAVR had a prior history of peripheral vascular disease (PVD) compared to ∼48–67% of those who underwent TAVR via alternative access. The details pertaining to individual access groups are available in **eTable-8, Data Supplement**.

**Table 1.** Baseline characteristics of patients and study details.

| Author(s) | Year | Region | Study Design | TAVR Groups | Sample Size | Mean age (yrs) | Males (%) | Diabetes (%) | Prior MI (%) | Prior PCI (%) | PVD (%) |
| --- | --- | --- | --- | --- | --- | --- | --- | --- | --- | --- | --- |
| Bleizziffer et al | 2009 | Germany | Single-center Retrospective cohort | TF/TA | 153/50 | 81.4/81.5 | 48/22 | - | - | 18/28 | 17/52 |
| Petronio et al | 2010 | Italy | Multicenter Prospective Cohort | TF/TSA | 460/54 | 83/83 | 41.3/66.7 | 27.4/20.4 | 20.7/33.3 | 27/46.3 | 15/55.6 |
| Ewe et al | 2011 | Singapore | Single-center Retrospective cohort | TF/TA | 45/59 | 82.2/79.4 | 46.7/52.5 | 29/27 | 22/24 | - | 11/67.8 |
| Taramasso et al | 2011 | Italy | Single-center Prospective Cohort | TF/TA/TSA | 140/16/19 | 79.8/78/79.7 | 53.1/31/73.7 | 18.6/25/26.3 | 22/37/36.8 | - | 18.6/62.5/63.1 |
| Eltchaninof et al | 2011 | France | Multicenter Prospective Cohort | TF/TA/TSA | 161/71/12 | 83/82/75.5 | 52.5/64.3/50 | 29.3/25.3/8.3 | 25.5/14/25 | - | - |
| Moynagh et al | 2011 | UK, Ireland | Multicenter Retrospective Cohort | TF/TSA | 253/35 | 81.7/80.6 | - | - | 16/34 | 24/34 | 21.3/74.2 |
| Petronio et al | 2012 | Italy | Retrospective Cohort | TF/TSA | 141/141 | 83/83 | 58/61 | - | - | - | 20.6/85.1 |
| Saia et al | 2012 | Italy | Single-center Prospective Cohort | TF/TSA/TA | 66/12/24 | - | - | - | - | - | - |
| Gillard et al | 2012 | France | Multicenter Prospective Cohort | TF/TA/TSA | 2361/567/184 | 83/81.5/82.2 | 47.4/58.6/71.2 | - | 14.5/25/18.5 | 15.2/30/24.2 | 12.5/48.1/41.6 |
| Muensterer et al | 2013 | Germany | Single-center Retrospective Cohort | TF/TSA | 301/40 | 80.2/79.5 | 44.9/57.5 | - | - | - | 14/42.5 |
| Lardizabal et al | 2013 | United States | Single-center Registry | TAO/TA | 44/76 | 85/87 | 43/53 | 23/25 | 18/24 | 20/50 | 73/75 |
| Adamo et al | 2014 | Italy | Single-center Prospective Cohort | TF/TAO/TSA | 170/44/32 | 83/83/82 | 62/61/44 | 28/34/28 | 17/16/16 | 24/25/41 | 11/70/66 |
| Blackman et al | 2014 | United Kingdom | Single-center Prospective Cohort | TF/TA/TSA | 1091/408/94 | 82/82/82 | 53/52/68 | 22/19.4/25 | 21.5/19.9/25.3 | - | 16.9/44/55.3 |
| Schymik et al | 2015 | Germany | Single-center Prospective Cohort | TF/TA | 587/413 | 82.1/81.5 | 39.7/49 | - | 58.3/64.4 | - | 10.9/22 |
| Frohllich et al | 2015 | United Kingdom | Multicenter Prospective Registry | TF/TA/TAO/TSA | 2828/761/185/188 | 83/84/82/83 | 51/56/49/65 | 23/22/21/23 | 22/22/23/28 | 21/20/18/24 | - |
| Koifman et al | 2016 | United States | Retrospective Cohort | TF/TA | 516/132 | 83/84 | 51/44 | 35/30 | - | 31/35 | 30/56 |
| Kirker et al | 2016 | United States | Retrospective Observational Cohort | TF/TA/TC | 100/12/25 | 83/82.5/77 | 51/42/52 | 34/50/48 | - | - | 39/83/80 |
| Arai et al | 2016 | France, Japan | Multicenter Prospective Cohort | TF/TAO/TA | 467/289/42 | 83.8/83.7/81.3 | 50/51/71 | 22/22/26 | 3/6/5 | - | 11/32/36 |
| Gleason et al | 2017 | United States | Post-hoc analysis | TF/TSA | 202/202 | 80.2/80.8 | 58.9/63.9 | 43/43 | 31.2/31.7 | 40/40 | 57.9/60.4 |
| Paone et al | 2018 | United | Retrospective Cohort | TF/TC/TCV | 373/32/58 | 80.4/79/79.6 | 55/50/44.8 | 41/34.4/44.8 | - | - | 23.3/78.1/75.9 |
| Watanabe et al | 2018 | Japan | Retrospective Cohort | TF/TC | 643/83 | 81.4/80 | 53.7/65.1 | 27/31.3 | 14.8/19.2 | 37.6/42.2 | 20.5/61.4 |
| van Wely et al | 2018 | Netherlands | Retrospective Cohort | TF/TSA | 29/91 | 82/80 | - | - | - | - | - |
| Dalmuji et al | 2018 | France, United States | Prospective Observational Cohort | TA/TAO/TSA/TC | 45/67/17/43 | 82/84/80/79.6 | 73/45/59/63 | 27/28/24/40 | 22/25/6/35 | - | - |
| Chollet et al | 2018 | France | Prospective Cohort | TF/TAO | 481/163 | 85/83.8 | 44/7/43.3 | 25.6/28.8 | - | - | 5/31.9 |
| Folliguet et al | 2019 | France | Prospective Observational Study | TF/TC | 10,598/435 | 83.2/81.1 | 47.3/59 | 25.7/31.1 | 37.9/46.4 | 28.9/38.4 | 17.3/50.7 |
| Amer et al | 2019 | United States | Single-center Retrospective Cohort | TSA/TC | 38/33 | 71/82 | 50/58 | 39/33 | 39/24 | 39/24 | 47/64 |
| Allen et al | 2019 | United States | Single-center Retrospective Cohort | TAO/TC/TAP | 33/84/48 | 78/78.9/76.8 | 45.5/54.8/50 | 36.4/54.8/35.4 | - | 42.4/61.9/39.6 | 36.4/73.8/50 |
| Long et al | 2020 | United States | Single-center Retrospective Cohort | SC/TCV | 41/16 | 83.2/80.7 | 36.4/41.4 | 43.9/31.8 | 12.2/27.3 | 9.8/13.6 | 29.3/36.4 |
| Leclercq et al | 2020 | France | Single-center Prospective Cohort | TF/TC | 51/81 | 81.8/81.6 | 33/69 | 42/36 | - | 33/52 | - |
| Hudziack et al | 2020 | Poland | Single-center Retrospective Cohort | TF/TC | 232/33 | 78.6/78 | 43.5/51.5 | 43/57 | 22/33 | 38/42.2 | 18/36.4 |
| Lu et al | 2020 | Switzerland | Single-center Retrospective Cohort | TF/TC | 255/51 | 83/83 | 49.8/60.8 | 24/27 | - | - | 14/41.2 |
| Kirker et al | 2020 | United States | Observational Registry Study | TSA/TC | 1576/788 | 78/78 | 52.8/52.3 | 41/41.9 | - | - | 73.2/73.2 |
| Jones et al | 2021 | United States | Single-center Retrospective Cohort | TF/TC | 1319/146 | 81/79 | 53/50 | 34/42 | - | - | 25/90 |
| Marie et al | 2021 | France | Single-center Retrospective Cohort | TF/TC | 400/100 | 81.3/79.9 | 50.5/69 | 26/27 | - | - | 14.8/86 |

Numerical values are represented as numbers (n) or percentages (%). TF: Transfemoral; TSA: Transaxillary/Subclavian; TA: Transapical; TAO: Transaortic; TC: Transcarotid; TCV: Transcaval; MI: Myocardial Infarction; PCI: Percutaneous Coronary Intervention; PVD: Peripheral Vascular Disease.

### Network Structure

**Figure 2** displays the network structure for the short-term and long-term risk of mortality associated with different access sites used in TAVR, with TF access being the point of reference. The overall network appears to be well-connected. There are a total 6 different access sites: TF, TSA, TAO, TA, TC, and TCV in the short-term and 5 (with the exception of TCV) in the long-term network models. Network structures for the respective secondary outcomes are shown in **eFigure-2, Data Supplement**.

**Figure 2.**
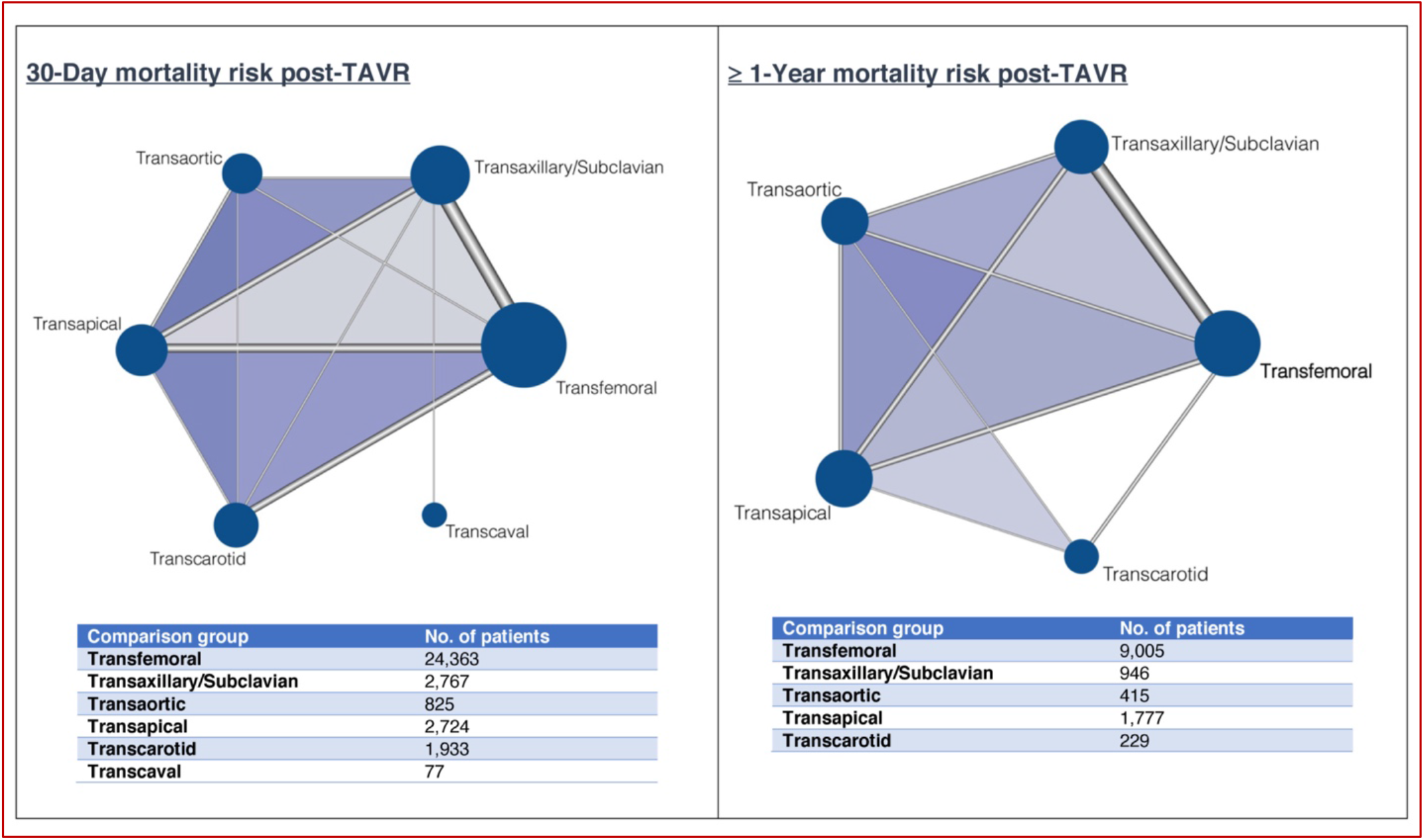
Network of primary outcomes: 30-days and ≥ 1-year mortality Each node (solid circle) represents an individual access site used in TAVR. The size of the node corresponds to the total number of patients included in each of the groups. The thickness of the edges (gray lines) connecting different nodes corresponds to the number of studies directly comparing a particular treatment pair.

### All-cause mortality

All 34 studies reported incidence of all-cause mortality at 30-days following TAVR. Compared with TF access, TAO access was associated with 87% risk of mortality [RR, 1.87 (95% CI, 1.38–2.54)]. A similar risk was observed with TA access [RR 1.89 (95% CI, 1.57–2.28)]. However, we did not find any difference in mortality with TSA [RR, 1.12 (95% CI, 0.85– 1.48)]; TC [RR, 1.19 (95% CI, 0.88–1.60)]; or TCV access [RR, 1.68 (95% CI, 0.46–6.10)] (**Figure 3**). A total of 12 studies (N = 12,372) reported all-cause mortality at ≥ 1-year following TAVR. The risk of all-cause mortality was significantly higher with TAO [RR, 1.35 (95% CI, 1.01–1.81)] or TA [RR, 1.44 (95% CI, 1.14–1.81)] access for TAVR, whereas the risk of mortality associated with TSA [RR, 1.16 (95% CI, 0.95–1.42)] and TC [RR, 0.92 (95% CI, 0.54–1.56)] access was comparable with TF access, as shown in **Figure 3**.

**Figure 3.**
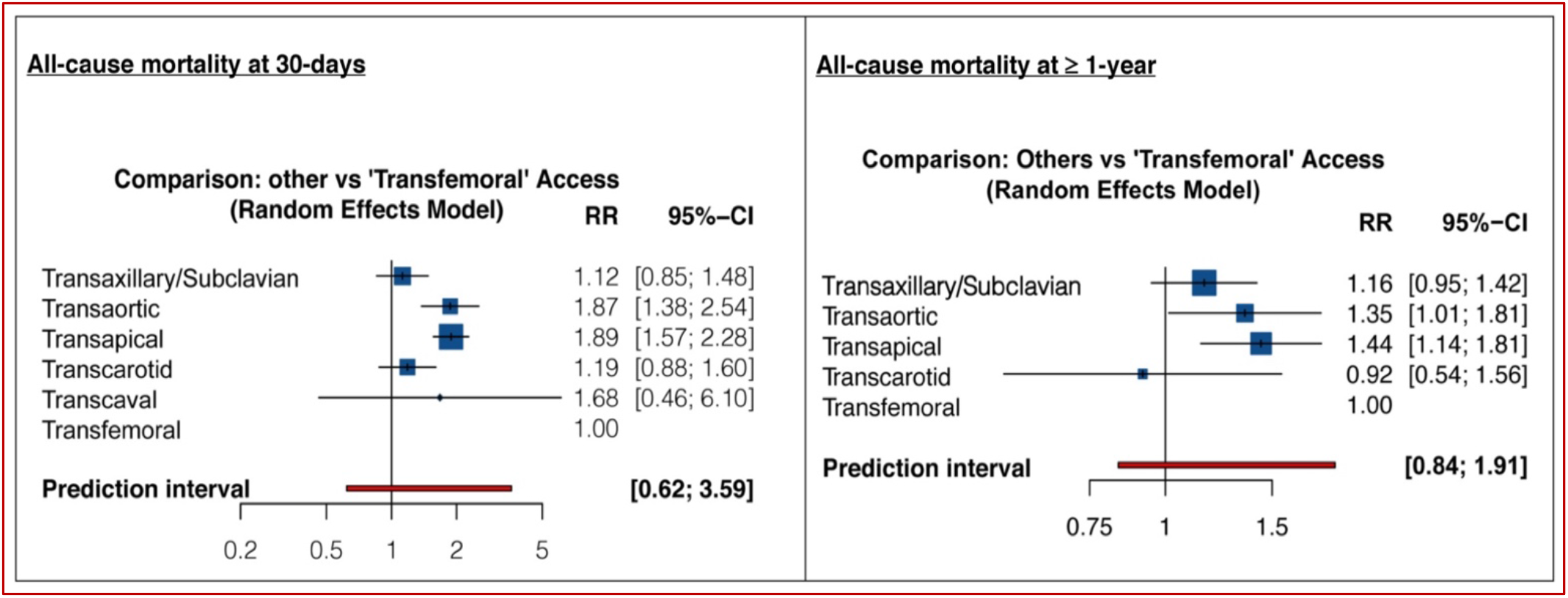
Forest Plots showing the RR (95% CI) for Primary Outcomes: 30-days and ≥ 1-year mortality.

### Conversion to open cardiac surgery

Eight studies (N = 15,833) reported the incidence of conversion of TAVR to open cardiac surgery. None of the alternative TAVR approaches were found to increase the predisposition to open cardiac surgery as compared to TF access — RR, 2.09 (95% CI, 0.59– 7.35) for TSA; RR, 1.95 (95% CI, 0.26–14.53)] for TAO; RR, 0.89 (95% CI, 0.11–7.39) for TA; and RR, 1.17 (95% CI 0.42–3.22) for TCV (**Figure 3**).

We identified a total of 26 studies (N = 28,333) that compared insertion of new pacemaker following TAVR across various access sites. Compared with TF access, both TAO [RR, 0.60 (95% CI, 0.40–0.89)] and TA access [RR, 0.49 (95%CI, 0.36–0.65)] significantly lowered the risk of new pacemaker insertion at 30-days of TAVR, whereas the risk with TSA or TC access was comparable with TF access. However, we observed that TCV access was associated with a 4.5-fold risk of pacemaker insertion [RR 4.45 (95% CI, 2.48–8.00)] as shown in **Figure 4**.

**Figure 4.**
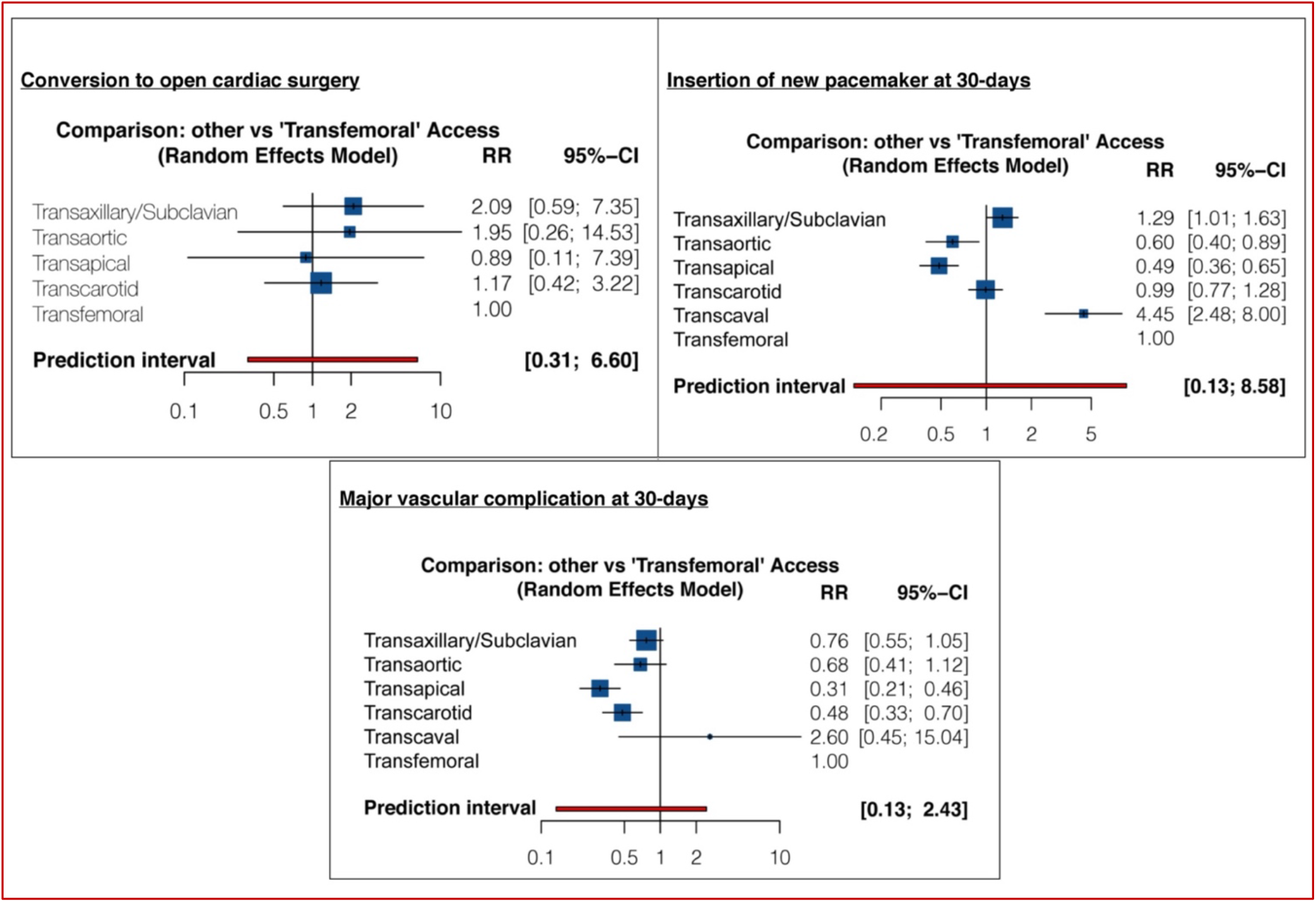
Forest Plots for Secondary Outcomes: conversion to cardiac surgery; insertion of new pacemaker at 30-days; and major vascular complications at 30-days.

### Major vascular complication at 30-days

Twenty-five studies with a pooled sample size of 26,024 patients reported the incidence of major vascular complications following TAVR. We found that patients who underwent TA [RR, 0.31 (95% CI, 0.21–0.46)] or TC TAVR [RR, 0.48 (95% CI, 0.33–0.70)] had a lower risk of developing major vascular complications within 30-days, as compared to patients who underwent TF TAVR. The risk profiles of TSA, TAO, or TCV access were similar to that of TF access (**Figure 4**).

### Stroke at 30-days

Compared with the TF access, there were no significant differences in stroke risk with TSA [RR, 1.45 (95% CI, 0.90–2.32)]; TAO [RR, 0.72 (95% CI, 0.33–1.55)]; TA [RR, 0.86 (95% CI, 0.48–1.55)]; TC [RR, 0.85 (95% CI, 0.53–1.38)]; and TCV [RR, 0.62 (95% CI, 0.07–5.25)] access for TAVR, as shown in **Figure 5**.

**Figure 5.**
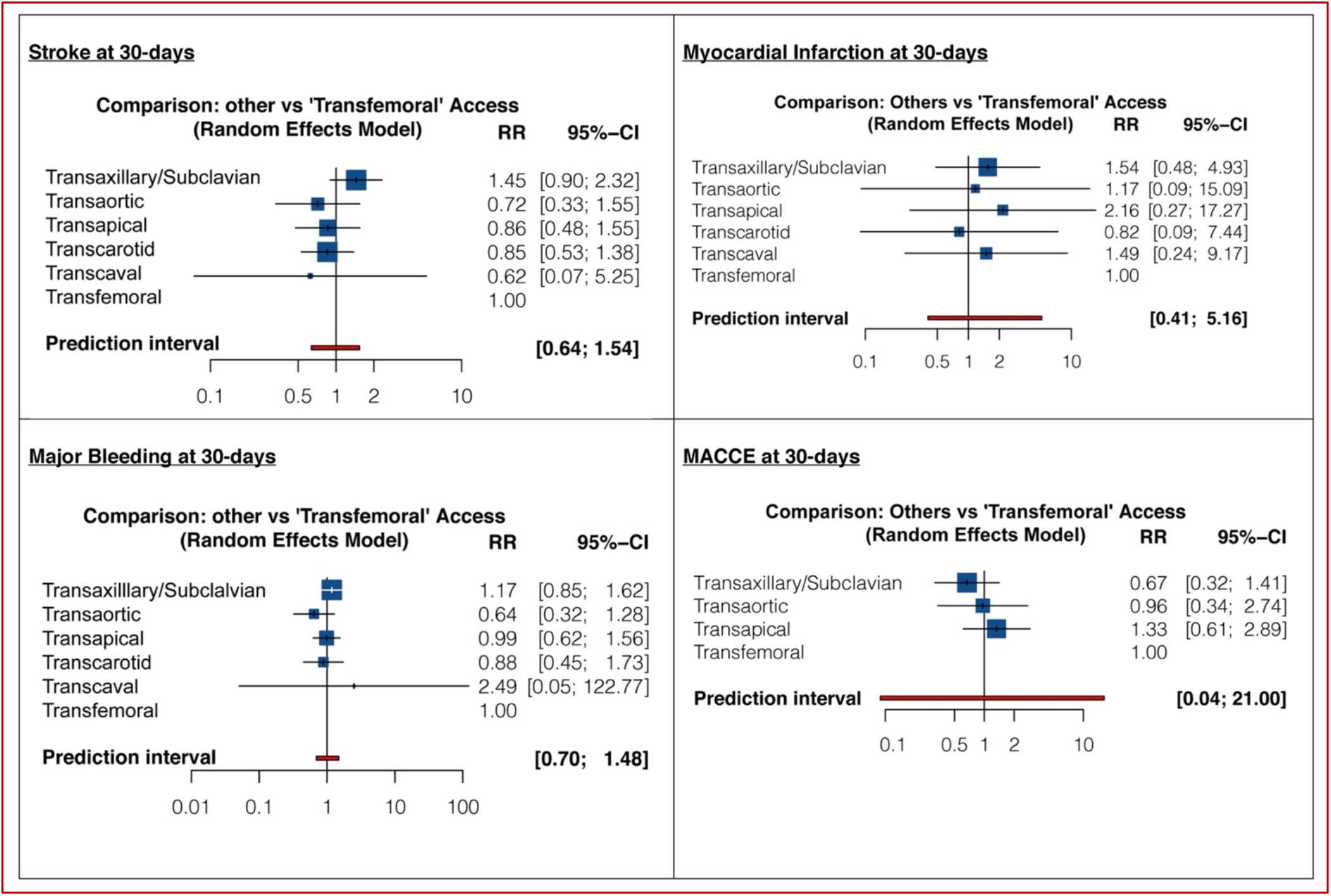
Forest plot for secondary outcomes: stroke at 30-days; myocardial infarction at 30-days; major bleeding at 30-days; and MACCE at 30-days.

### Myocardial Infarction at 30-days

Nine studies with a pooled patient population of 3,330 were identified for comparing the risk of myocardial infarction occurring within 30-days post-TAVR across 6 different access sites. When compared with the TF access, none of the other access sites were found to have a significantly different risk of myocardial infarction at 30-days (**Figure 5**).

### Major bleeding at 30-days

We identified 10 studies (N = 4,546) reporting the incidence of major bleeding among their patient cohorts at 30-days post-TAVR. There were no significant differences in the risk of major bleeding with TSA [RR, 1.17 (95% CI, 0.85–1.62)]; TAO [RR, 0.64 (95% CI, 0.32–1.28)]; TA [RR, 0.99 (95% CI, 0.62–1.56)]; TC [RR, 0.88 (95% CI, 0.45–1.73)]; and TCV [RR, 2.49 (95% CI, 0.05–122.77)] access as compared to TF access (**Figure 5**).

### MACCE at 30-days

Only 5 studies (N = 1,199) evaluated the risk of MACCE at 30-days across 4 different TAVR access sites. Compared to TF access, none of the other access sites had a significantly different risk of MACCE with an RR of 0.67 (95% CI, 0.32– 1.41) for TSA; RR, 0.96 (95% CI, 0.34–2.74) for TAO; and RR 1.33 (95% CI, 0.61–2.89) for TA, as illustrated in **Figure 5**.

### Ranking Strategies for TAVR Access Site

Transfemoral approach was ranked the best strategy (P-score, 0.89), followed by TSA (P-score, 0.71) and TC (P-score, 0.64) for reducing all-cause mortality at 30-days, whereas both the TAO (P-score, 0.19) and TA (P-score, 0.18) approaches were relatively least effective in reducing the risk of mortality at 30-days post-TAVR (**Figure 6**). At 1-year follow-up, TF access was ranked the most effective (P-score, 0.82), followed by TC (P-score, 0.81) for limiting all-cause mortality risk at ≥ 1-year (**Figure 6**). Rankograms for the secondary outcomes are shown in **eFigure-3, Data Supplement**.

**Figure 6.**
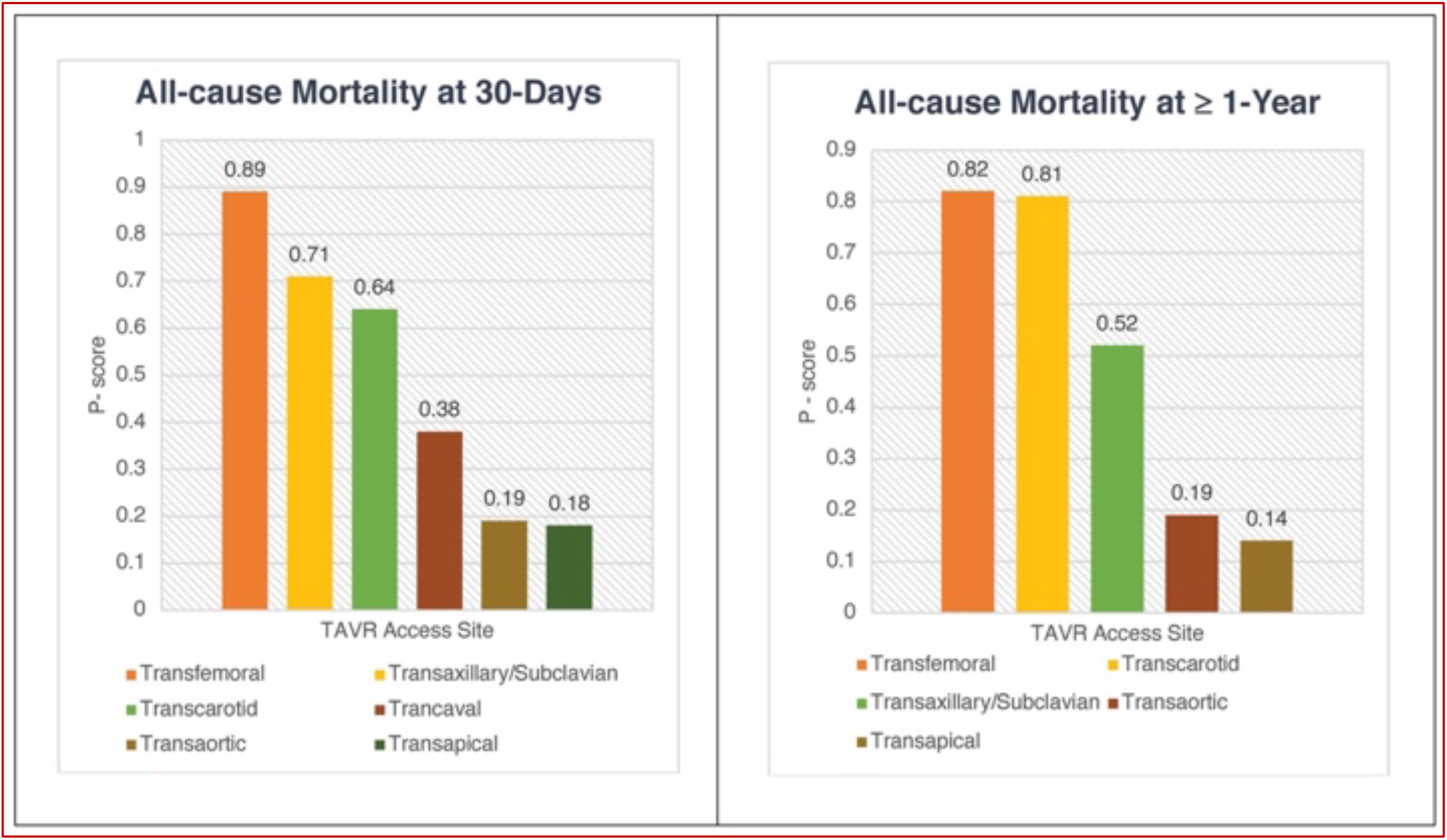
Rankograms for Primary Outcomes.

## Discussion

This network meta-analysis including 34 studies with 30,986 patients provides a cross-comparison of various available access sites and attempts to generate a pooled estimate using the currently available evidence. Multiple observations of this study are noteworthy and deserve emphasis — 1) As compared to TF access, both TAO and TA access were associated with a nearly two-fold increase in all-cause mortality at 30-days and ≥ 1-year following TAVR; 2) Major adverse cardiovascular and cerebrovascular events were comparable across various TAVR access sites; 3) Non-thoracic alternative access sites (TC, TSA, and TCV) were associated with comparable mortality and similar risk of stroke as compared to TF access; 4) TA and TC access for TAVR were associated with lower risk of vascular complications as compared to TF access; 5) TCV access for TAVR was associated with an increased risk of permanent pacemaker implant as compared to TF access.

The TF route is currently considered the gold standard for TAVR because TAVR devices were primarily designed for TF approach. Among the few initially available approaches for TAVR (including TF, TA, and TAO), the TF route was considered the least invasive one, not requiring a surgical incision. Since then, centers are using TF as their default approach.^5,54^ Thanks to ongoing development of transcatheter heart valve technologies, including progressive evolution and downsizing of delivery systems and introducer sheaths, TF TAVR is now feasible in the majority of patients. However, despite the technical advancements in valve delivery systems and reduced sheath sizes, a small but non-negligible group of patients comprising 10-15% of cases are still not suitable for TF access due to tortuous, calcified, or small caliber iliofemoral arteries warranting alternative access TAVR approaches.^31,34^ In early TAVR experience, TAO and TA accesses were the only alternative access routes for valve implantation, followed by TSA, TC, and TCV. These approaches are limited by relative contraindications, such as respiratory failure in case of TA, and porcelain aorta and previous heart surgery, in cases of TAO. As a result, and with the downsizing of delivery systems for newer generation devices, interest in TA and TAO approaches waned and non-thoracic alternative access routes gained popularity.^30,36^ Prior studies have found both TAO and TA approaches similar to TF in terms of mortality-related outcomes.^32,47,55^ Our findings, on the contrary, indicate otherwise — underscoring nearly a two-fold risk of mortality with TAO and TA approaches compared with TF access both in short as well as long-term. The increased mortality with TA and TAO approaches has been ascribed to high “risk profiles” of patients and a steeper learning curve for the procedure.^7,30,31,56^ This is in line with findings from a prior study of high-risk patients from the FRANCE-2 TAVI Registry demonstrating a higher 1-year adjusted mortality (HR 1.45; 95% CI, 1.09–1.92) with TA as compared to TF TAVR.^7^

Stroke has been well-recognized as a dreaded complication of TAVR, associated with a five-fold increase in 30-day risk of mortality.^57^ In this network meta-analysis, we observed that 30-day risk of stroke was comparable across different approaches, with no particular approach having a higher or lower predisposition to stroke. While some early studies of TC TAVR suggested a higher than ideal stroke rate, our analysis corroborates contemporary findings from the French registry where the stroke rate was similar to that of TF TAVR^36,58^ and lower than what has been reported for TSA TAVR.^56^

Though TCV and the other peripheral alternative access had comparable outcomes compared to TF access, there was a significant increased risk of permanent pacemaker with TCV compared to TF access. No information on non-modifiable causes (pre-existing conduction disorders, distribution and amount of calcification, left ventricular outflow geometry) and mechanical factors of the procedure (implantation depth, transcatheter valve oversizing, and radial force of the valve at outflow level) are available to look for association of this findings. More data analysis comparing TCV and TF access including the above-mentioned factors are needed to explore this finding.

Our findings highlight that alternative access options that avoid the thoracic cavity, including TC, TSA and TCV access, best mimic the safety profile of TF TAVR. Of these, TC demonstrated the most favorable outcomes, with a lower number of vascular complications as compared to TCV, TSA, and TF access, and similar reduction in risk of 1-year mortality as compared to TF access. Probable reasons for this could be the superficial location of the carotid arteries and the comfort level of cardiovascular surgeons with carotid artery manipulation, facilitating access and closure under direct visualization, potentially reducing major or life-threatening bleeding and vascular complications.^59^ However, there are currently no randomized data to support any alternative access strategy over another so the strategy should be chosen by the heart team based on patient anatomy, risk factors, and the center’s expertise. While further studies comparing TC, to TCV or TSA are needed to inform which access site should preferred when alternative access is needed, several recent publications suggest reduced stroke and improved 30-day outcomes with TC compared to TSA and TAO/TA.^16,34,38^

Despite our network meta-analysis results and outcomes from single or larger multicenter experiences,^60^ the selection of the optimal access route — including TF — should always be individualized to the patient, operator and center experience. Additionally, to preserve the benefit of TF access, some centers have adopted intravascular lithotripsy, in the femoral and iliac vasculature prior to TAVR in patients with inadequate femoral access to decrease the need for alternative access.^61^ There are currently no data to support the use of peripheral vascular interventions to obtain femoral access versus pursuing an alternative access strategy in a carotid or axillary artery with minimal atherosclerosis. TAVR computed tomography should be routinely performed with imaging of the complete peripheral vascular anatomy. This should be reviewed by the multidisciplinary heart team including cardiology, cardiothoracic surgery, and radiology to determine the preferred access strategy prior to the case.

## Limitations

The present study has several limitations. First, the inclusion of all study designs instead of just randomized studies could result in the introduction of bias at the individual study level. Second, the baseline characteristics, unequal sample sizes, definitions of clinical outcomes, and differences in the follow-up durations may add to imprecision. The overall incidence of events was low, which could have influenced our inability to reach statistically significant results for some of the outcome measures. We also lacked ability to adjust for peri-procedural characteristics (prosthetic valve size/type, depth of implantation, valve-in-valve, native valve morphology), which may significantly influence the outcomes. Furthermore, the antithrombotic regimen and role of embolic protection devices used may have contributed as possible confounders. Finally, indirect comparisons in network meta-analysis are built on the assumption of transitivity, implying that studies making different direct comparisons must be sufficiently similar in all respects. Although this approach respects randomization, it does not represent randomized evidence.

## Conclusion

Non-thoracic alternative access routes (TC, TSA and TCV) are associated with a similar safety profile as compared to TF access with regarding the major complications of periprocedural mortality, stroke, and major bleeding following TAVR, while TA and TAO access are associated with increased short- and long-term mortality.

## Data Availability

All the data used in this manuscript has been extracted from already published articles. The authors declare that all supporting data are available within the article (and its online supplementary files).

## Acknowledgements

None.

## CRediT statement for author contribution

**SR:** Conceptualization, Methodology, Writing – Review & Editing, Validation; **SL:** Conceptualization, Software, Formal analysis, Data curation, Methodology, Writing – Original Draft, Visualization; **KA:** Validation, Writing – Review & Editing, Methodology; **AKC:** Validation, Writing – Review & Editing, **SV:** Data curation, Resources, Writing - Writing – Review & Editing; **PV:** Validation, Writing – Review & Editing, Supervision, Funding acquisition.

