## Supplementary material for "Network Meta-Analysis Comparing the Short and Long-Term Outcomes of Alternative Access for Transcatheter Aortic Valve Replacement": Data Supplement

eTable 1. PRISMA Checklist reporting necessary items.

| Section/topic | # | Checklist item | Reported on page # |
|---|---|---|---|
| **TITLE** | | | |
| Title | 1 | Identify the report as a systematic review, meta-analysis, or both. | 1 |
| **ABSTRACT** | | | |
| Structured summary | 2 | Provide a structured summary including, as applicable: background; objectives; data sources; study eligibility criteria, participants, and interventions; study appraisal and synthesis methods; results; limitations; conclusions and implications of key findings; systematic review registration number. | 2 |
| **INTRODUCTION** | | | |
| Rationale | 3 | Describe the rationale for the review in the context of what is already known. | 3 |
| Objectives | 4 | Provide an explicit statement of questions being addressed with reference to participants, interventions, comparisons, outcomes, and study design (PICOS). | 3 |
| **METHODS** | | | |
| Protocol and registration | 5 | Indicate if a review protocol exists, if and where it can be accessed (e.g., Web address), and, if available, provide registration information including registration number. | - |
| Eligibility criteria | 6 | Specify study characteristics (e.g., PICOS, length of follow-up) and report characteristics (e.g., years considered, language, publication status) used as criteria for eligibility, giving rationale. | 5 |
| Information sources | 7 | Describe all information sources (e.g., databases with dates of coverage, contact with study authors to identify additional studies) in the search and date last searched. | 4 |
| Search | 8 | Present full electronic search strategy for at least one database, including any limits used, such that it could be repeated. | 4 |
| Study selection | 9 | State the process for selecting studies (i.e., screening, eligibility, included in systematic review, and, if applicable, included in the meta-analysis). | 5 |
| Data collection process | 10 | Describe method of data extraction from reports (e.g., piloted forms, independently, in duplicate) and any processes for obtaining and confirming data from investigators. | 5 |
| Data items | 11 | List and define all variables for which data were sought (e.g., PICOS, funding sources) and any assumptions and simplifications made. | 5 |
| Risk of bias in individual studies | 12 | Describe methods used for assessing risk of bias of individual studies (including specification of whether this was done at the study or outcome level), and how this information is to be used in any data synthesis. | 5 |
| Summary measures | 13 | State the principal summary measures (e.g., risk ratio, difference in means). | 6 |
| Synthesis of results | 14 | Describe the methods of handling data and combining results of studies, if done, including measures of consistency (e.g., $I^2$) for each meta-analysis. | 6 – 7 |

| Section/topic | # | Checklist item | Reported on page # |
|---|---|---|---|
| Risk of bias across studies | 15 | Specify any assessment of risk of bias that may affect the cumulative evidence (e.g., publication bias, selective reporting within studies). | 6 |
| Additional analyses | 16 | Describe methods of additional analyses (e.g., sensitivity or subgroup analyses, meta-regression), if done, indicating which were pre-specified. | Data Supplement |
| **RESULTS** | | | |
| Study selection | 17 | Give numbers of studies screened, assessed for eligibility, and included in the review, with reasons for exclusions at each stage, ideally with a flow diagram. | 6 -7 |
| Study characteristics | 18 | For each study, present characteristics for which data were extracted (e.g., study size, PICOS, follow-up period) and provide the citations. | 6 -7 |
| Risk of bias within studies | 19 | Present data on risk of bias of each study and, if available, any outcome level assessment (see item 12). | Data Supplement |
| Results of individual studies | 20 | For all outcomes considered (benefits or harms), present, for each study: (a) simple summary data for each intervention group (b) effect estimates and confidence intervals, ideally with a forest plot. | 6 – 10 |
| Synthesis of results | 21 | Present results of each meta-analysis done, including confidence intervals and measures of consistency. | 6 – 10 |
| Risk of bias across studies | 22 | Present results of any assessment of risk of bias across studies (see Item 15). | Data Supplement |
| Additional analysis | 23 | Give results of additional analyses, if done (e.g., sensitivity or subgroup analyses, meta-regression [see Item 16]). | Data Supplement |
| **DISCUSSION** | | | |
| Summary of evidence | 24 | Summarize the main findings including the strength of evidence for each main outcome; consider their relevance to key groups (e.g., healthcare providers, users, and policy makers). | 11 – 13 |
| Limitations | 25 | Discuss limitations at study and outcome level (e.g., risk of bias), and at review-level (e.g., incomplete retrieval of identified research, reporting bias). | 13 - 14 |
| Conclusions | 26 | Provide a general interpretation of the results in the context of other evidence, and implications for future research. | 14 |
| **FUNDING** | | | |
| Funding | 27 | Describe sources of funding for the systematic review and other support (e.g., supply of data); role of funders for the systematic review. | 14 |

*From:* Moher D, Liberati A, Tetzlaff J, Altman DG, The PRISMA Group (2009). Preferred Reporting Items for Systematic Reviews and Meta-Analyses: The PRISMA Statement. PLoS Med 6(7): e1000097. doi:10.1371/journal.pmed1000097

For more information, visit: **www.prisma-statement.org**.

eTable 2. Number of articles yielded by our search hits on PubMed/Medline.

| No. | Search terms | Records |
|---|---|---|
| 1. | (transcatheter aortic valve replacement) AND (transfemoral) | 1476 |
| 2. | (transcatheter aortic valve replacement) AND (transaortic) | 298 |
| 3. | ((clinical outcomes) OR (mortality)) AND (transcatheter aortic) | 6548 |
| 4. | (tavr) AND (access site) | 134 |
| 5. | (transcatheter aortic valve) AND ((subclavian) OR (transaxillary)) | 285 |
| 6. | (transaortic transcatheter*) AND (cardiovascular outcomes) | 210 |
| 7. | ((((transapical) OR (transfemoral)) OR (subclavian)) AND (transcatheter aortic*)) NOT (surgical) | 313 |

eTable 3. Number of articles yielded by our search hits on Cochrane library.

| No. | Search terms | Records |
|---|---|---|
| 1. | “transcatheter aortic valve” AND “mortality” NOT “surgical” | 236 |
| 2. | "transcatheter aortic valve" AND “transfemoral” OR “transaortic” | 269 |
| 3. | “transapical” AND “access site” OR “tavr” | 465 |
| 4. | "transcatheter aortic" AND “cardiovascular outcomes” | 03 |
| 5. | “transcatheter aortic valve implantation” AND “versus” NOT “surgical” | 101 |
| 6. | “transcatheter aortic valve implantation” AND “subclavian” OR “transcaval” OR “transaxillary” | 58 |

eTable 4. Quality appraisal of studies included in this network meta-analysis using MINORS criteria.

| | Clearly stated aim | Inclusion of consecutive patients | Prospective collection of data | Endpoints appropriate to the aim of the study. | Unbiased assessment of study endpoint | Follow-up period appropriate to the aim of the study | Loss to follow-up < 5% | Prospective calculation of the study size | An adequate control group | Contemporary groups | Baseline equivalence of groups | Adequate statistical analyses | Total score |
|---|---|---|---|---|---|---|---|---|---|---|---|---|---|
| **Study author (year)** | | | | | | | | | | | | | |
| Petronio et al (2010) | 2 | 2 | 2 | 2 | 0 | 2 | 2 | 2 | 2 | 2 | 2 | 2 | **22** |
| Taramasso et al (2011) | 2 | 2 | 2 | 2 | 0 | 2 | 2 | 2 | 2 | 2 | 2 | 2 | **22** |
| Eltchaninof et al (2011) | 2 | 0 | 2 | 2 | 0 | 2 | 2 | 2 | 2 | 2 | 0 | 2 | **18** |
| Moynagh et al (2011) | 2 | 2 | 2 | 2 | 0 | 2 | 2 | 1 | 2 | 2 | 2 | 2 | **21** |
| Petronio et al (2012) | 2 | 2 | 2 | 2 | 0 | 2 | 2 | 2 | 2 | 2 | 2 | 2 | **22** |
| Saia et al (2012) | 2 | 0 | 2 | 2 | 0 | 2 | 2 | 2 | 2 | 2 | 0 | 2 | **18** |
| Gillard et al (2012) | 2 | 2 | 2 | 2 | 0 | 2 | 2 | 2 | 2 | 2 | 2 | 2 | **22** |
| Muensterer et al (2013) | 2 | 0 | 2 | 2 | 0 | 2 | 2 | 2 | 2 | 2 | 2 | 2 | **20** |
| Lardizabal et al (2013) | 2 | 2 | 2 | 2 | 0 | 2 | 2 | 2 | 0 | 2 | 2 | 2 | **20** |
| Adamo et al (2014) | 2 | 0 | 2 | 2 | 0 | 2 | 2 | 2 | 2 | 2 | 2 | 2 | **20** |
| Blackman et al (2014) | 2 | 2 | 2 | 2 | 0 | 2 | 0 | 2 | 2 | 2 | 2 | 2 | **20** |
| Frohilch et al (2015) | 2 | 0 | 2 | 2 | 0 | 2 | 1 | 2 | 2 | 2 | 2 | 2 | **19** |
| Koifman et al (2016) | 2 | 1 | 2 | 2 | 0 | 2 | 0 | 2 | 2 | 2 | 2 | 2 | **19** |
| Kirker et al (2016) | 2 | 0 | 2 | 2 | 0 | 2 | 2 | 2 | 2 | 2 | 2 | 2 | **20** |
| Arai et al (2016) | 2 | 1 | 2 | 2 | 0 | 2 | 0 | 2 | 2 | 2 | 0 | 2 | **17** |
| Gleason et al (2017) | 2 | 0 | 2 | 2 | 0 | 2 | 0 | 2 | 2 | 2 | 2 | 2 | **18** |
| Paone et al (2018) | 2 | 0 | 2 | 2 | 0 | 2 | 0 | 2 | 2 | 2 | 2 | 2 | **18** |
| Watanabe et al (2018) | 2 | 0 | 2 | 2 | 0 | 2 | 0 | 2 | 2 | 2 | 2 | 2 | **19** |
| Van Wely et al (2018) | 2 | 0 | 2 | 2 | 0 | 2 | 0 | 1 | 2 | 2 | 2 | 2 | **17** |

| | | | | | | | | | | | | |
|---|---|---|---|---|---|---|---|---|---|---|---|---|
| Dalmuji et al (2018) | 2 | 1 | 2 | 2 | 0 | 2 | 0 | 2 | 0 | 2 | 2 | 2 | **17** |
| Chollet el al (2018) | 2 | 2 | 2 | 2 | 0 | 2 | 0 | 2 | 2 | 2 | 2 | 2 | **20** |
| Folliguet et al (2019) | 2 | 2 | 2 | 2 | 0 | 2 | 0 | 2 | 2 | 2 | 0 | 2 | **19** |
| Amer et al (2019) | 2 | 1 | 2 | 2 | 0 | 2 | 0 | 2 | 0 | 2 | 2 | 2 | **17** |
| Allen et al (2020) | 2 | 2 | 2 | 2 | 0 | 2 | 2 | 2 | 0 | 2 | 2 | 2 | **20** |
| Long et al (2020) | 2 | 2 | 2 | 2 | 0 | 2 | 2 | 2 | 0 | 2 | 2 | 2 | **20** |
| Lu et al (2020) | 2 | 2 | 2 | 2 | 0 | 2 | 0 | 2 | 2 | 2 | 2 | 2 | **20** |
| Kirker et al (2020) | 2 | 0 | 2 | 2 | 0 | 2 | 0 | 2 | 0 | 2 | 2 | 2 | **16** |
| Jones et al (2021) | 2 | 1 | 2 | 2 | 0 | 2 | 0 | 2 | 2 | 2 | 0 | 2 | **17** |
| Marie et al (2021) | 2 | 1 | 2 | 2 | 0 | 2 | 0 | 2 | 2 | 2 | 2 | 2 | **19** |

The items are score 0 (not reported), 1 (reported but inadequate), or 2 (reported and adequate); ideal score being 24.

**eTable 5**. Consistency Evaluation of Network Meta-analysis Model and Separate Direct and Indirect Evidence (SIDE) Using Back-Calculation Method.

| Comparison | k | prop | NMA | Direct | Indirect | ROR | z | p-value* |
|---|---|---|---|---|---|---|---|---|
| **30-Day All-cause Mortality** | | | | | | | | |
| TAx/SC: Transaortic | 3 | 0.24 | 0.601 | 0.412 | 0.676 | 0.610 | -1.051 | 0.293 |
| TAx/SC: Transapical | 7 | 0.55 | 0.596 | 0.537 | 0.676 | 0.793 | -0.70 | 0.459 |
| TAx/SC: Transcarotid | 3 | 0.46 | 0.948 | 1.216 | 0.768 | 1.583 | 1.310 | 0.190 |
| TAx/SC: Transcaval | 1 | 0.17 | 0.670 | 1.410 | 0.576 | 2.449 | 0.504 | 0.614 |
| TAx/SC: Transfemoral | 13 | 0.77 | 1.125 | 1.036 | 1.490 | 0.695 | -1.085 | 0.277 |
| Transaortic: Transapical | 5 | 0.63 | 0.991 | 0.959 | 1.047 | 0.917 | -0.257 | 0.797 |
| Transaortic: Transcarotid | 2 | 0.21 | 1.576 | 2.671 | 1.373 | 1.946 | 1.309 | 0.190 |
| Transaortic: Transcaval | 0 | 0 | 1.115 | . | 1.115 | . | . | . |
| Transaortic: Transfemoral | 4 | 0.71 | 1.870 | 1.703 | 2.340 | 0.728 | -0.925 | 0.355 |
| Transapical: Transcarotid | 3 | 0.13 | 1.591 | 1.475 | 1.609 | 0.916 | -0.169 | 0.866 |
| Transapical: Transcaval | 0 | 0 | 1.125 | . | 1.125 | . | . | . |
| Transapical: Transfemoral | 12 | 0.92 | 1.888 | 1.918 | 1.559 | 1.230 | 0.577 | 0.564 |
| Transcarotid: Transcaval | 1 | 0.19 | 0.707 | 0.360 | 0.829 | 0.434 | -0.486 | 0.627 |
| Transcarotid: Transfemoral | 9 | 0.63 | 1.186 | 1.500 | 0.795 | 1.887 | 1.989 | 0.467 |
| Transcaval: Transfemoral | 1 | 0.82 | 1.678 | 1.878 | 0.992 | 1.893 | 0.369 | 0.712 |
| **1-Year All-cause Mortality** | | | | | | | | |
| TAx/SC: Transaortic | 2 | 0.66 | 0.814 | 1.038 | 0.507 | 2.048 | 1.816 | 0.069 |
| TAx/SC: Transapical | 3 | 0.74 | 0.804 | 0.825 | 0.747 | 1.104 | 0.319 | 0.749 |
| TAx/SC: Transcarotid | 0 | 0 | 1.267 | . | 1.267 | . | . | . |
| TAx/SC: Transfemoral | 8 | 1.00 | 1.167 | 1.154 | 433.478 | 0.003 | -2.450 | 0.014 |
| Transaortic: Transapical | 2 | 0.69 | 0.989 | 1.185 | 0.662 | 1.790 | 1.452 | 0.146 |
| Transaortic: Transcarotid | 1 | 0.17 | 1.557 | 10.761 | 1.055 | 10.197 | 2.757 | 0.005 |
| Transaortic: Transfemoral | 2 | 0.78 | 1.434 | 1.227 | 2.482 | 0.494 | -1.660 | 0.096 |
| Transapical: Transcarotid | 1 | 0.13 | 1.575 | 4.024 | 1.371 | 2.936 | 1.237 | 0.217 |
| Transapical: Transfemoral | 3 | 0.88 | 1.451 | 1.587 | 0.770 | 2.060 | 1.977 | 0.048 |
| Transcarotid: Transfemoral | 2 | 0.87 | 0.921 | 1.170 | 0.180 | 6.509 | 2.302 | 0.021 |
| **Conversion to Open Cardiac Surgery** | | | | | | | | |
| TAx/SC: Transaortic | 1 | 0.40 | 1.067 | 0.771 | 1.327 | 0.518 | -0.275 | 0.783 |
| TAx/SC: Transapical | 1 | 0.41 | 2.350 | 0.867 | 4.688 | 0.185 | -0.806 | 0.419 |
| TAx/SC: Transcarotid | 2 | 0.83 | 1.784 | 1.971 | 1.089 | 1.809 | 0.439 | 0.660 |
| TAx/SC: Transfemoral | 2 | 0.35 | 2.085 | 1.415 | 2.561 | 0.553 | -0.439 | 0.660 |
| Transaortic: Transapical | 2 | 1.00 | 2.203 | 2.203 | . | . | . | . |
| Transaortic: Transcarotid | 1 | 0.79 | 1.672 | 1.074 | 8.466 | 0.127 | -0.942 | 0.346 |
| Transaortic: Transfemoral | 0 | 0 | 1.955 | . | 1.955 | . | . | . |
| Transapical: Transcarotid | 1 | 0.72 | 0.759 | 0.956 | 0.417 | 2.294 | 0.385 | 0.699 |
| Transapical: Transfemoral | 0 | 0 | 0.887 | . | 0.887 | . | . | . |
| Transcarotid: Transfemoral | 3 | 0.82 | 1.169 | 1.299 | 0.718 | 1.809 | 0.439 | 0.661 |
| **30-Day New Pacemaker** | | | | | | | | |
| TAx/SC: Transaortic | 1 | 0.33 | 2.153 | 3.255 | 1.759 | 1.851 | 1.255 | 0.290 |
| TAx/SC: Transapical | 4 | 0.58 | 2.644 | 3.745 | 1.641 | 2.283 | 2.324 | 0.020 |

| | | | | | | | | |
|---|---|---|---|---|---|---|---|---|
| TAx/SC: Transcarotid | 2 | 0.33 | 1.297 | 1.125 | 1.393 | 0.807 | -0.634 | 0.526 |
| TAx/SC: Transcaval | 1 | 0.22 | 0.289 | 1.390 | 0.185 | 7.535 | 2.687 | 0.007 |
| TAx/SC: Transfemoral | 9 | 0.79 | 1.285 | 1.181 | 1.776 | 0.665 | -1.372 | 0.170 |
| Transaortic: Transapical | 3 | 0.52 | 1.228 | 1.264 | 1.190 | 1.062 | 0.126 | 0.899 |
| Transaortic: Transcarotid | 0 | 0 | 0.602 | . | 0.602 | . | . | . |
| Transaortic: Transcaval | 0 | 0 | 0.134 | . | 0.134 | . | . | . |
| Transaortic: Transfemoral | 3 | 0.92 | 0.597 | 0.634 | 0.309 | 2.055 | 0.969 | 0.332 |
| Transapical: Transcarotid | 0 | 0 | 0.491 | . | 0.491 | . | . | . |
| Transapical: Transcaval | 0 | 0 | 0.109 | . | 0.109 | . | . | . |
| Transapical: Transfemoral | 8 | 0.93 | 0.486 | 0.509 | 0.267 | 1.902 | 1.098 | 0.272 |
| Transcarotid: Transcaval | 1 | 0.18 | 0.223 | 0.121 | 0.256 | 0.473 | -0.907 | 0.364 |
| Transcarotid: Transfemoral | 8 | 0.81 | 0.991 | 0.961 | 1.136 | 0.846 | -0.494 | 0.621 |
| Transcaval: Transfemoral | 1 | 0.75 | 0.449 | 6.431 | 1.444 | 4.454 | 2.153 | 0.031 |
| **30-Day Major Vascular Complication** | | | | | | | | |
| TAx/SC: Transaortic | 3 | 0.35 | 1.123 | 0.774 | 1.368 | 0.566 | -0.931 | 0.352 |
| TAx/SC: Transapical | 5 | 0.48 | 2.449 | 2.260 | 2.633 | 0.858 | -0.311 | 0.755 |
| TAx/SC: Transcarotid | 3 | 0.43 | 1.587 | 1.334 | 1.811 | 0.737 | -0.685 | 0.493 |
| TAx/SC: Transcaval | 1 | 0.21 | 0.294 | 0.470 | 0.260 | 1.810 | 0.265 | 0.790 |
| TAx/SC: Transfemoral | 11 | 0.81 | 0.763 | 0.831 | 0.524 | 1.586 | 1.099 | 0.271 |
| Transaortic: Transapical | 3 | 0.54 | 2.181 | 1.622 | 3.072 | 0.528 | -1.097 | 0.272 |
| Transaortic: Transcarotid | 1 | 0.17 | 1.413 | 1.676 | 1.363 | 1.229 | 0.254 | 0.799 |
| Transaortic: Transcaval | 0 | 0 | 0.261 | . | 0.261 | . | . | . |
| Transaortic: Transfemoral | 3 | 0.71 | 0.679 | 0.897 | 0.344 | 2.610 | 1.719 | 0.085 |
| Transapical: Transcarotid | 1 | 0.13 | 0.648 | 2.103 | 0.546 | 3.855 | 1.678 | 0.093 |
| Transapical: Transcaval | 0 | 0 | 0.120 | . | 0.120 | . | . | . |
| Transapical: Transfemoral | 5 | 0.84 | 0.311 | 0.245 | 1.099 | 0.223 | -2.781 | 0.005 |
| Transcarotid: Transcaval | 1 | 0.31 | 0.185 | 0.600 | 0.108 | 5.539 | 0.872 | 0.383 |
| Transcarotid: Transfemoral | 7 | 0.69 | 0.481 | 0.474 | 0.495 | 0.958 | -0.102 | 0.918 |
| Transcaval: Transfemoral | 1 | 0.75 | 2.598 | 3.831 | 0.825 | 4.643 | 0.745 | 0.456 |
| **30-Day Stroke** | | | | | | | | |
| TAx/SC: Transaortic | 2 | 0.18 | 2.019 | 0.896 | 2.416 | 0.371 | -0.882 | 0.378 |
| TAx/SC: Transapical | 3 | 0.15 | 1.684 | 0.827 | 1.909 | 0.433 | -0.825 | 0.409 |
| TAx/SC: Transcarotid | 3 | 0.85 | 1.694 | 1.781 | 1.262 | 1.411 | 0.687 | 0.492 |
| TAx/SC: Transcaval | 1 | 1.00 | 2.317 | 2.371 | . | . | . | . |
| TAx/SC: Transfemoral | 7 | 0.58 | 1.446 | 1.301 | 1.873 | 0.778 | -0.512 | 0.608 |
| Transaortic: Transapical | 3 | 0.53 | 0.834 | 0.912 | 0.754 | 1.209 | 0.221 | 0.825 |
| Transaortic: Transcarotid | 2 | 0.33 | 0.839 | 1.410 | 0.651 | 2.165 | 0.844 | 0.398 |
| Transaortic: Transcaval | 0 | 0 | 1.147 | . | 1.147 | . | . | . |
| Transaortic: Transfemoral | 2 | 0.54 | 0.716 | 0.527 | 1.025 | 0.514 | 0.844 | 0.398 |
| Transapical: Transcarotid | 3 | 0.26 | 1.006 | 1.614 | 0.850 | 1.899 | 0.786 | 0.431 |
| Transapical: Transcaval | 0 | 0 | 1.376 | . | 1.376 | . | . | . |
| Transapical: Transfemoral | 5 | 0.75 | 0.859 | 0.805 | 1.044 | 0.771 | -0.373 | 0.709 |
| Transcarotid: Transcaval | 0 | 0 | 1.368 | . | 1.368 | . | . | . |
| Transcarotid: Transfemoral | 6 | 0.45 | 0.853 | 1.250 | 0.628 | 1.991 | 1.395 | 0.163 |
| Transcaval: Transfemoral | 0 | 0 | 0.624 | . | 0.624 | . | . | . |

| | | | | | | | | |
|---|---|---|---|---|---|---|---|---|
| **30-Day MI** | | | | | | | | |
| TAx/SC: Transaortic | 1 | 0.46 | 1.318 | 1.369 | 1.275 | 1.074 | 0.026 | 0.979 |
| TAx/SC: Transapical | 1 | 0.34 | 0.713 | 1.960 | 0.423 | 4.632 | 0.632 | 0.527 |
| TAx/SC: Transcarotid | 0 | 0 | 1.889 | . | 1.889 | . | . | . |
| TAx/SC: Transcaval | 1 | 1.00 | 1.034 | 1.034 | . | . | . | . |
| TAx/SC: Transfemoral | 5 | 0.99 | 1.543 | 1.544 | 1.415 | 1.091 | 0.151 | 0.988 |
| Transaortic: Transapical | 1 | 0.65 | 0.541 | 0.344 | 1.262 | 0.272 | -0.499 | 0.617 |
| Transaortic: Transcarotid | 0 | 0 | 1.434 | . | 1.434 | . | . | . |
| Transaortic: Transcaval | 0 | 0 | 0.785 | . | 0.785 | . | . | . |
| Transaortic: Transfemoral | 1 | 0.43 | 1.171 | 3831 | 0.482 | 7.954 | 0.786 | 0.431 |
| Transapical: Transcarotid | 1 | 0.40 | 2.649 | 2.040 | 3.420 | 0.596 | -0.186 | 0.852 |
| Transapical: Transcaval | 0 | 0 | 1.449 | . | 1.449 | . | . | . |
| Transapical: Transfemoral | 2 | 0.72 | 2.163 | 2.170 | 2.146 | 1.011 | 0.004 | 0.996 |
| Transcarotid: Transcaval | 0 | 0 | 0.547 | . | 0.547 | . | . | . |
| Transcarotid: Transfemoral | 2 | 0.93 | 0.816 | 0.923 | 0.158 | 5.840 | 0.398 | 0.690 |
| Transcaval: Transfemoral | 0 | 0 | 1.492 | . | 1.492 | . | . | . |
| **30-Day Major Bleeding** | | | | | | | | |
| TAx/SC: Transaortic | 0 | 0 | 1.824 | . | 1.824 | . | . | . |
| TAx/SC: Transapical | 0 | 0 | 1.189 | . | 1.189 | . | . | . |
| TAx/SC: Transcarotid | 1 | 0.14 | 1.329 | 0.522 | 1.543 | 0.338 | -1.001 | 0.317 |
| TAx/SC: Transcaval | 1 | 1.00 | 0.470 | 0.470 | . | . | . | . |
| TAx/SC: Transfemoral | 2 | 0.98 | 1.172 | 1.203 | 0.407 | 2.955 | 1.001 | 0.317 |
| Transaortic: Transapical | 1 | 0.61 | 0.652 | 0.440 | 1.103 | 0.366 | -1.433 | 0.152 |
| Transaortic: Transcarotid | 0 | 0 | 0.729 | . | 0.729 | . | . | . |
| Transaortic: Transcaval | 0 | 0 | 0.258 | . | 0.258 | . | . | . |
| Transaortic: Transfemoral | 1 | 0.52 | 0.643 | 1.045 | 0.382 | 2.736 | 1.433 | 0.152 |
| Transapical: Transcarotid | 1 | 0.04 | 1.118 | 2.040 | 1.088 | 1.876 | 0.312 | 0.755 |
| Transapical: Transcaval | 0 | 0 | 0.395 | . | 0.395 | . | . | . |
| Transapical: Transfemoral | 2 | 0.87 | 0.986 | 0.869 | 2.339 | 0.372 | -1.416 | 0.156 |
| Transcarotid: Transcaval | 0 | 0 | 0.354 | . | 0.354 | . | . | . |
| Transcarotid: Transfemoral | 3 | 0.88 | 0.882 | 0.789 | 2.044 | 0.386 | -0.889 | 0.374 |
| Transcaval: Transfemoral | 0 | 0 | 2.494 | . | 2.494 | . | . | . |
| **30-Day MACCE** | | | | | | | | |
| TAx/SC: Transaortic | 0 | 0 | 0.693 | . | 0.693 | . | . | . |
| TAx/SC: Transapical | 2 | 0.69 | 0.503 | 0.695 | 0.248 | 2.803 | 0.927 | 0.354 |
| TAx/SC: Transfemoral | 4 | 1.00 | 0.668 | 0.673 | 0.118 | 5.706 | 0.297 | 0.766 |
| Transaortic: Transapical | 1 | 1.00 | 0.726 | 0.726 | . | . | . | . |
| Transaortic: Transfemoral | 0 | 0 | 0.964 | . | 0.964 | . | . | . |
| Transapical: Transfemoral | 2 | 0.99 | 1.328 | 1.413 | 0.010 | 142.392 | 1.3829 | 0.166 |

**k:** Number of studies providing direct evidence; **Prop:** direct evidence proportion; **NMA:** Estimated treatment effect (RR) in the network meta-analysis; **Direct:** Estimated treatment effect (RR) derived from direct evidence; **Indirect:** Estimated treatment effect (RR) derived from indirect evidence; **ROR:** Ratio of ratios (direct vs indirect); **z:** z-value of test for disagreement (direct vs indirect); **P-value**: p-value of test for disagreement/inconsistency (direct vs indirect); **TAX/SC:** Transaxillary/Subclavian.
*p<0.05 indicates a significant disagreement (inconsistency) between direct and indirect estimate.

**eTable-6.** Assessment of heterogeneity (Q) and inconsistency among models comparing primary and secondary outcomes.

| Outcomes | $I^2$ | $\tau^2$ | Q within design (p-value)* | Q between design (p-value)* | Total Q (p-value)* |
|---|---|---|---|---|---|
| 30-Day All-cause mortality | 0 | 0 | 13.57 (0.808) | 15.60 (0.210) | 29.17 (0.560) |
| ≥ 1-year All-cause mortality | 55.9 | 0.04 | 2.47 (0.871) | 29.28 (0.0003) | 31.76 (0.004) |
| Conversion to cardiac surgery | 0 | 0 | 1.48 (0.686) | 1.35 (0.716) | 2.84 (0.829) |
| 30-Day New Pacemaker | 52.5 | 0.08 | 21.39 (0.06) | 29.12 (0.002) | 50.51 (0.001) |
| 30-Day Vascular Complication | 0 | 0 | 9.44 (0.738) | 12.06 (0.522) | 21.51 (0.715) |
| 30-Day Stroke | 0 | 0 | 1.45 (0.993) | 5.01 (0.957) | 6.46 (0.998) |
| 30-Day MI | 0 | 0 | 0.03 (0.986) | 1.90 (0.863) | 1.92 (0.963) |
| 30-Day Major Bleeding | 0 | 0 | 0.09 (0.765) | 0.36 (0.835) | 0.45 (0.929) |
| 30-Day MACCE | 2.2 | 0.012 | 1.18 (0.757) | 2.91 (0.080) | 4.09 (0.393) |

*p-value <0.10 suggests a significant heterogeneity is present. Total Q represents heterogeneity within and between designs.

**eTable-7**. Pairwise comparisons in the network meta-analyses for the entire cohort.

**1. All-cause Mortality at 30-Days**

| | | | | | |
|---|---|---|---|---|---|
| **Tax/SC** | 0.41 (0.18 – 0.92) | 0.54 (0.36 – 0.81) | 1.22 (0.73 – 2.02) | 1.41 (0.06 – 33.65) | 1.04 (0.76 – 1.42) |
| 0.60 (0.41 – 0.89) | **Transaortic** | 0.96 (0.64 – 1.43) | 2.67 (1.10 – 6.48) | . | 1.70 (1.18 – 2.45) |
| 0.60 (0.44 – 0.81) | 0.99 (0.72 – 1.36) | **Transapical** | 1.47 (0.57 – 3.80) | . | 1.92 (1.58 – 2.33) |
| 0.95 (0.67 – 1.33) | 1.58 (1.05 – 2.36) | 1.59 (1.13 – 2.24) | **Transcarotid** | 0.36 (0.02 – 7.41) | 1.50 (1.03 – 2.19) |
| 0.67 (0.18 – 2.48) | 1.11 (0.30 – 4.20) | 1.13 (0.31 – 4.14) | 0.71 (0.19 – 2.65) | **Transcaval** | 1.88 (0.45 – 7.79) |
| 1.12 (0.85 – 1.48) | 1.87 (1.38 – 2.54) | 1.89 (1.57 – 2.28) | 1.19 (0.88 – 1.60) | 1.68 (0.46 – 6.10) | **Transfemoral** |

**2. All-cause Mortality at ≥ 1-Year**

| | | | | |
|---|---|---|---|---|
| **Tax/SC** | 1.04 (0.66 – 1.63) | 0.82 (0.61 – 1.12) | . | 1.15 (0.94 – 1.42) |
| 0.81 (0.56 – 1.17) | **Transaortic** | 1.18 (0.76 – 1.84) | 10.76 (2.39 – 48.51) | 1.23 (0.83 – 1.82) |
| 0.80 (0.62 – 1.05 | 0.99 (0.69 – 1.42) | **Transapical** | 4.02 (0.81 – 19.90) | 1.59 (1.23 – 2.04) |
| 1.27 (0.72 – 2.23) | 1.56 (0.84 – 2.88) | 1.57 (0.89 – 2.80) | **Transcarotid** | 1.17 (0.66 – 2.07) |
| 1.17 (0.95 – 1.43) | 1.43 (1.02 – 2.03) | 1.45 (1.15 – 1.84) | 0.92 (0.54 – 1.57) | **Transfemoral** |

**3. Conversion to open cardiac surgery**

| | | | | |
|---|---|---|---|---|
| **Tax/SC** | 0.77 (0.04 – 15.35) | 0.87 (0.04 – 20.28) | 1.97 (0.67 – 5.84) | 1.42 (0.17 – 12.03) |
| 1.07 (0.16 – 7.11) | **Transaortic** | 2.20 (0.56 – 8.63) | 1.07 (0.15 – 7.84) | . |
| 2.35 (0.31 – 17.66) | 2.20 (0.56 – 8.63) | **Transapical** | .0.96 (0.10 – 8.84) | . |
| 1.78 (0.66 – 4.80) | 1.67 (0.29 – 9.74) | 0.76 (0.11 – 5.03) | **Transcarotid** | 1.30 (0.43 – 3.97) |
| 2.09 (0.59 – 7.35) | 1.95 (0.26 – 14.53) | 0.89 (0.11 – 7.39) | 1.17 (0.42 – 3.22) | **Transfemoral** |

**4. Major Vascular Complication at 30-Days**

| | | | | | |
|---|---|---|---|---|---|
| **Tax/SC** | 0.77 (0.29 – 2.04) | 2.26 (1.13 – 4.53) | 1.33 (0.69 – 2.58) | 0.47 (0.01–23.13) | 0.83 (0.58 – 1.19) |
| 1.12 (0.63 – 1.99) | **Transaortic** | 1.62 (0.75 – 3.53) | 1.68 (0.40 – 7.10) | . | 0.90 (0.52 – 1.62) |
| 2.45 (1.52 – 3.96) | 2.18 (1.23 – 3.85) | **Transapical** | 2.10 (0.48 – 9.17) | . | 0.25 (0.16 – 0.37) |
| 1.59 (1.03 – 2.45) | 1.41 (0.77 – 2.59) | 0.65 (0.38 – 1.10) | **Transcarotid** | 0.60 (0.02 – 14.55) | 0.47 (0.30 – 0.75) |
| 0.29 (0.05 – 1.72) | 0.26 (0.04 – 1.62) | 0.12 (0.02 – 0.72) | 0.18 (0.03 – 1.10) | **Transcaval** | 3.83 (0.50 – 29.21) |
| 0.76 (0.55 – 1.05) | 0.68 (0.41 – 1.12) | 0.31 (0.21 – 0.46) | 0.48 (0.33 – 0.70) | 2.60 (0.45 – 15.04) | **Transfemoral** |

**5. New Pacemaker at 30-Days**

| | | | | | |
|---|---|---|---|---|---|
| **Tax/SC** | 3.25 (1.48 – 7.15) | 3.75 (2.38 – 5.89) | 1.12 (0.66 – 1.93) | 1.39 (0.38 – 5.10) | 1.18 (0.91 – 1.54) |
| 2.15 (1.37 – 3.38) | **Transaortic** | 1.26 (0.66 – 2.42) | . | . | 0.63 (0.42 – 0.97) |
| 2.64 (1.87 – 3.73) | 1.34 (0.82–2.19) | **Transapical** | . | . | 0.46 (0.38 – 0.69) |
| 1.30 (0.95 – 1.77) | 0.61 (0.37–1.01) | 0.49 (0.33 – 0.72) | **Transcarotid** | 0.12 (0.03 – 0.52) | 0.91 (0.72 – 1.28) |
| 0.29 (0.16 – 0.53) | 0.13 (0.06–0.28) | 0.11 (0.06 – 0.21) | 0.22 (0.12 – 0.42) | **Transcaval** | 6.43 (3.27 – 12.64) |
| 1.29 (1.01 – 1.63) | 0.58 (0.39–0.88) | 0.49 (0.36 – 0.65) | 0.99 (0.77 – 1.28) | 4.45 (2.48 – 8.00) | **Transfemoral** |

**6. Stroke at 30-Days**

| | | | | | |
|---|---|---|---|---|---|
| **TAx/SC** | 0.90 (0.12 – 6.59) | 0.83 (0.13–5.17) | 1.78 (1.23 – 2.59) | 2.32 (0.29 – 18.49) | 1.30 (0.70 – 2.42) |
| 2.02 (0.86 – 4.72) | **Transaortic** | 0.19 (0.29 – 2.89) | 1.41 (0.32 – 6.13) | . | 0.53 (0.18 – 1.50) |
| 1.68 (0.83 – 3.42) | 0.83 (0.36 – 1.93) | **Transapical** | 1.61 (0.41 – 6.36) | . | 0.80 (0.41 – 1.59) |
| 1.69 (1.20 – 2.40) | 0.84 (0.36 – 1.95) | 1.01 (0.50 – 2.03) | **Transcarotid** | . | 1.25 (0.61 – 2.57) |
| 2.32 (0.29 – 18.49) | 1.15 (0.36 – 1.93) | 1.38 (0.15 – 12.35) | 1.37 (0.17 – 11.23) | **Transcaval** | . |
| 1.45 (0.90 – 2.32) | 0.72 (0.33 – 1.55) | 0.86 (0.48 – 1.55) | 0.85 (0.53 – 1.38) | 0.62 (0.07 – 5.25) | **Transfemoral** |

**7. Myocardial Infarction at 30-Days**

| | | | | | |
|---|---|---|---|---|---|
| **TAx/SC** | 1.37 (0.03 – 67.22) | 1.96 (0.04 – 93.06) | . | 1.03 (0.26 – 4.17) | 1.54 (0.48 – 4.97) |
| 1.32 (0.09 – 18.40) | **Transaortic** | 0.34 (0.02 – 7.00) | . | . | 3.83 (0.08 – 190.43) |
| 0.71 (0.07 – 6.79) | 0.54 (0.05 – 6.16) | **Transapical** | 2.04 (0.04 – 96.93) | . | 2.17 (0.19 – 25.20) |
| 1.89 (0.16 – 22.36) | 1.43 (0.06 – 35.90) | 2.65 (0.18 – 40.02) | **Transcarotid** | . | 0.92 (0.09 – 9.12) |
| 1.03 (0.26 – 4.17) | 0.78 (0.04 – 15.49) | 1.45 (0.10 – 20.51) | 0.55 (0.03 – 9.34) | **Transcaval** | . |
| 1.54 (0.48 – 4.93) | 1.17 (0.09 – 15.09) | 2.16 (0.27 – 17.27) | 0.82 (0.09 – 7.44) | 1.49 (0.24 – 9.17) | **Transfemoral** |

**8. Major Bleeding at 30-Days**

| | | | | | |
|---|---|---|---|---|---|
| **TAx/SC** | . | . | 0.52 (0.07 – 3.75) | 0.47 (0.01 – 22.82) | 1.20 (0.87 – 1.67) |
| 1.82 (0.85 – 3.90) | **Transaortic** | 0.44 (0.19 – 1.04) | . | . | 1.04 (0.40 – 2.72) |
| 1.19 (0.68 – 2.08) | 0.65 (0.19 – 1.04) | **Transapical** | 2.04 (0.04 – 96.93) | . | 0.87 (0.53 – 1.42) |
| 1.33 (0.64 – 2.76) | 0.73 (0.03 – 22.45) | 1.12 (0.50 – 2.52) | **Transcarotid** | . | 0.79 (0.39 – 1.62) |
| 0.47 (0.01 – 22.82) | 0.26 (0.00 – 71.37) | 0.40 (0.01 – 19.98) | 0.35 (0.01 – 18.38) | **Transcaval** | . |
| 1.17 (0.85 – 1.62) | 0.64 (0.05 – 28.27) | 0.99 (0.62 – 1.56) | 0.88 (0.45 – 1.73) | 2.49 (0.05 – 122.77) | **Transfemoral** |

**9. MACCE at 30-Days**

| | | | |
|---|---|---|---|
| **TAx/SC** | . | 0.69 (0.21 – 2.35) | 0.67 (0.32 – 1.43) |
| 0.69 (0.20 – 2.37) | **Transaortic** | 0.73 (0.36 – 1.46) | . |
| 0.50 (0.18 – 1.38) | 0.73 (0.36 – 1.46) | **Transapical** | 1.41 (0.65 – 65.09) |
| 0.67 (0.32 – 1.41) | 0.96 (0.34 – 2.74) | 1.33 (0.61 – 2.89) | **Transfemoral** |

**eTable 8**. Baseline Characteristics of Studies and Patients in Different Comparison Groups.

| **Characteristics** | **Transfemoral** | **Transaxillary/Subclavian** | **Transaortic** | **Transapical** | **Transcarotid** | **Transcaval** |
|---|---|---|---|---|---|---|
| No. of studies (patients) | 28 (24,363) | 17 (2,767) | 7 (825) | 15 (2,724) | 13 (1,933) | 2 (77) |
| Age, years (mean ± SD) | 81.9 ± 7.1 | 80.5 ± 6.9 | 82.9 ± 16.1 | 81.6 ± 8.1 | 79.7 ± 8.4 | 80.1 ± 6.8 |
| Males (%) | 49.5 | 58.4 | 48.2 | 51.4 | 58.0 | 43.1 |
| Diabetes Mellitus (%) | 30.0 | 29.2 | 27.6 | 28.4 | 38.7 | 38.3 |
| Prior MI (%) | 23.2 | 25.5 | 17.6 | 25.8 | 31.5 | 27.3 |
| Prior PCI (%) | 28.1 | 32.3 | 26.3 | 33.7 | 43.5 | 13.6 |
| Prior PVD (%) | 22.1 | 52.4 | 48.6 | 54.2 | 66.8 | 56.1 |
| NYHA > III or IV (%) | 71.1 | 68.0 | 78.6 | 73.3 | 56.3 | 94.0 |
| STS Risk | 8.7 ± 5.4 | 12.0 ± 13.1 | 7.8 ± 3.8 | 11.7 ± 6.3 | 7.6 ± 4.8 | 8.5 ± 3.5 |

**Fgure-1.** Graphical representation of the inconsistency among primary and secondary outcomes.

**a. All-cause Mortality At 30-Days**

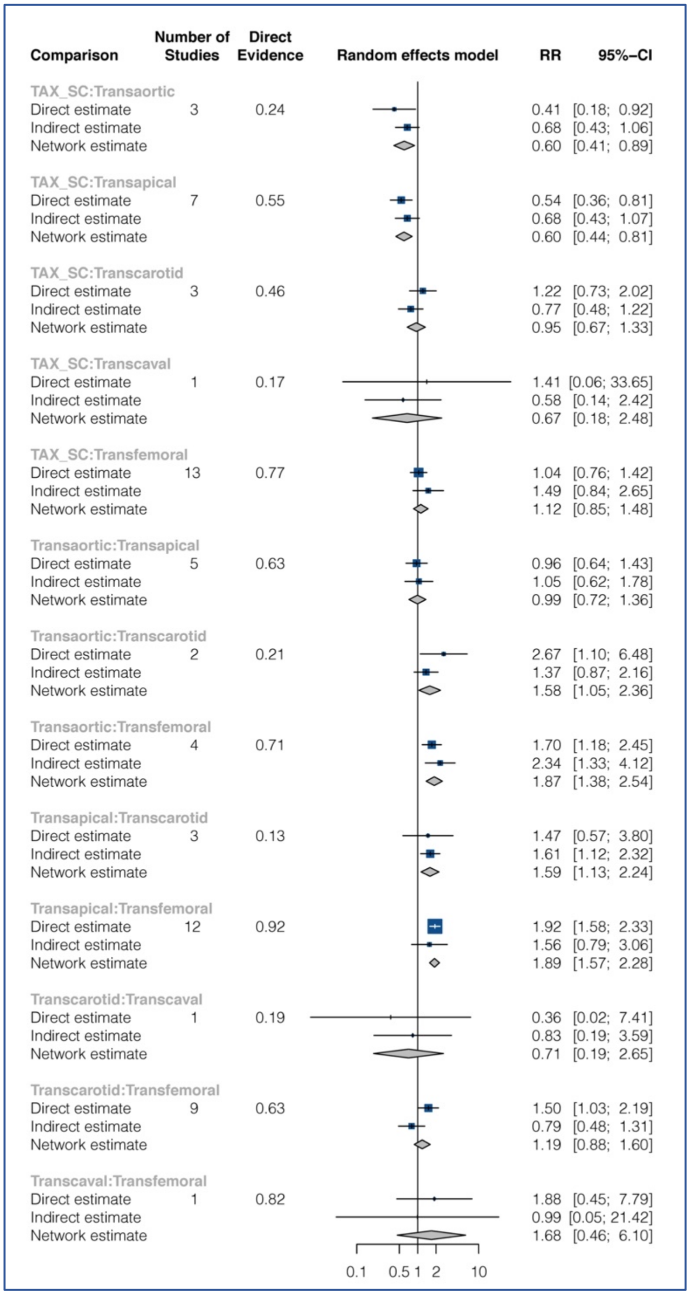


**b. All-cause Mortality at ≥1-Year**

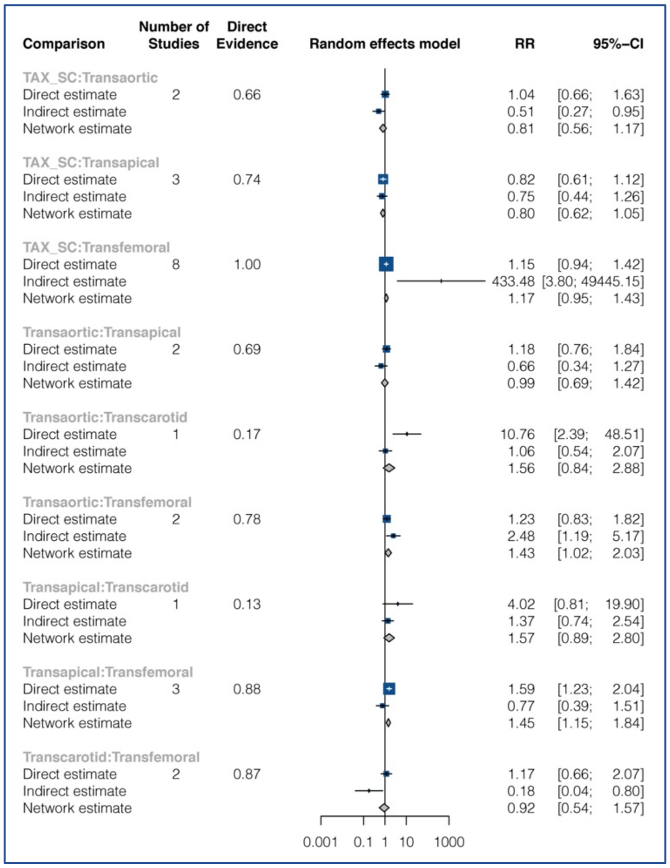

**c. Conversion to open cardiac surgery**

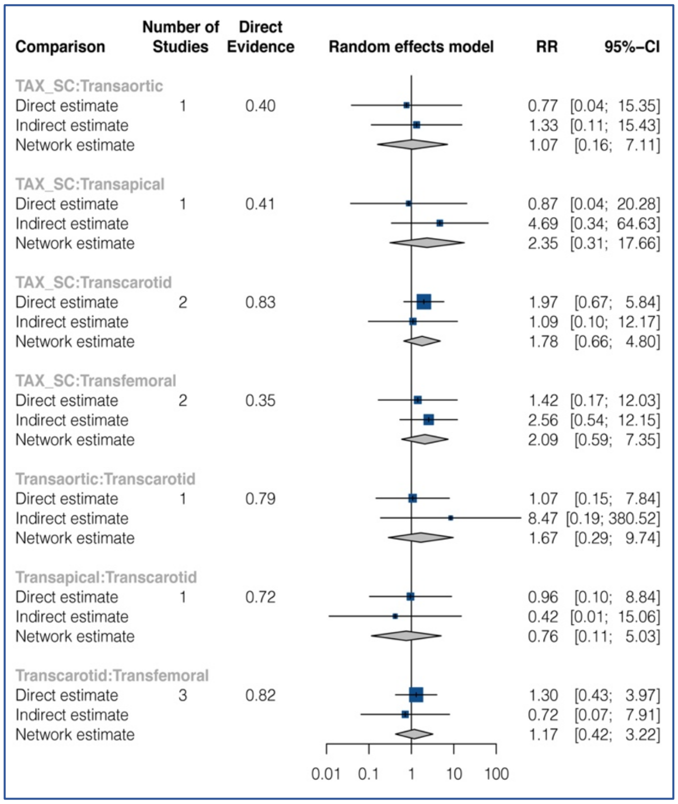


**d. New pacemaker insertion at 30-days**

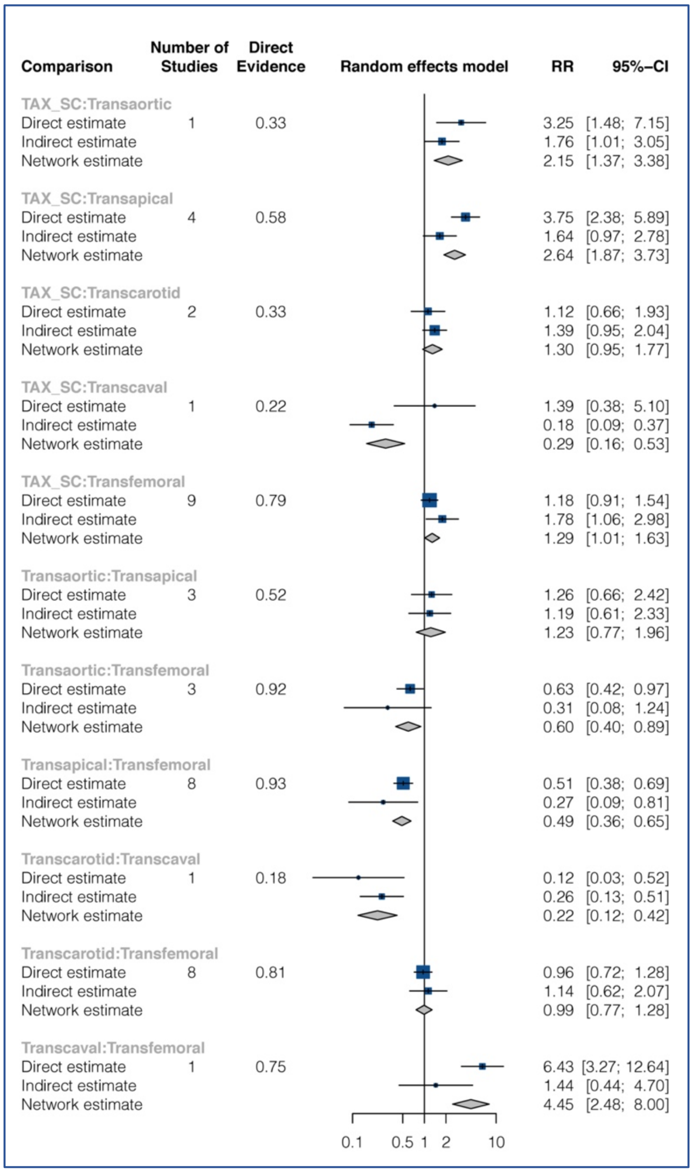

### e. Major vascular complication at 30-days.

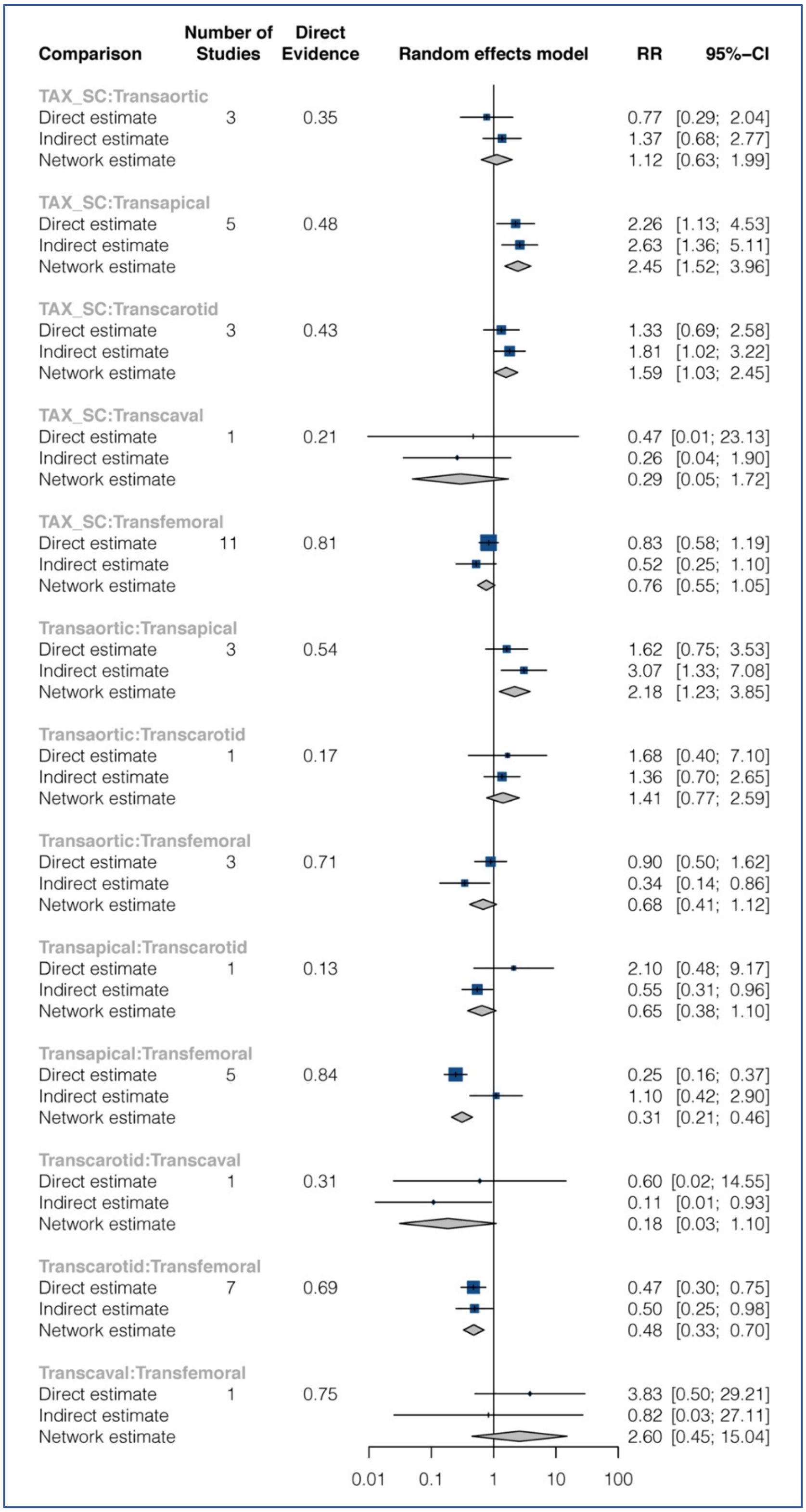

**f. Stroke At 30-Days**

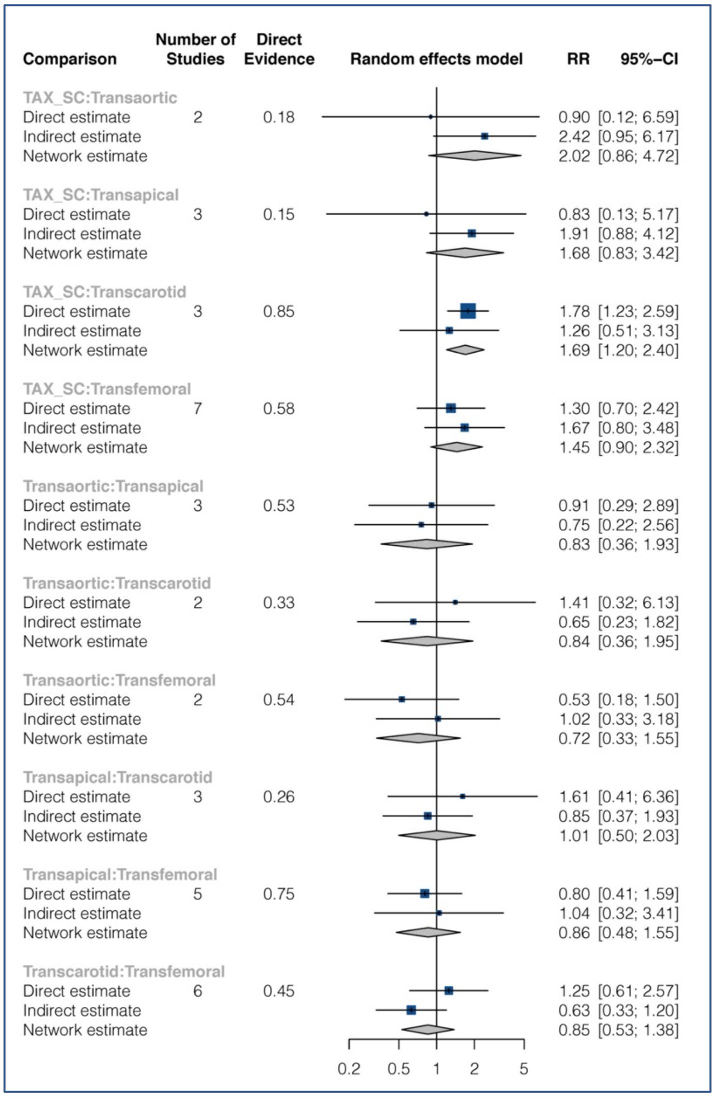


**g. MI At 30-Days**

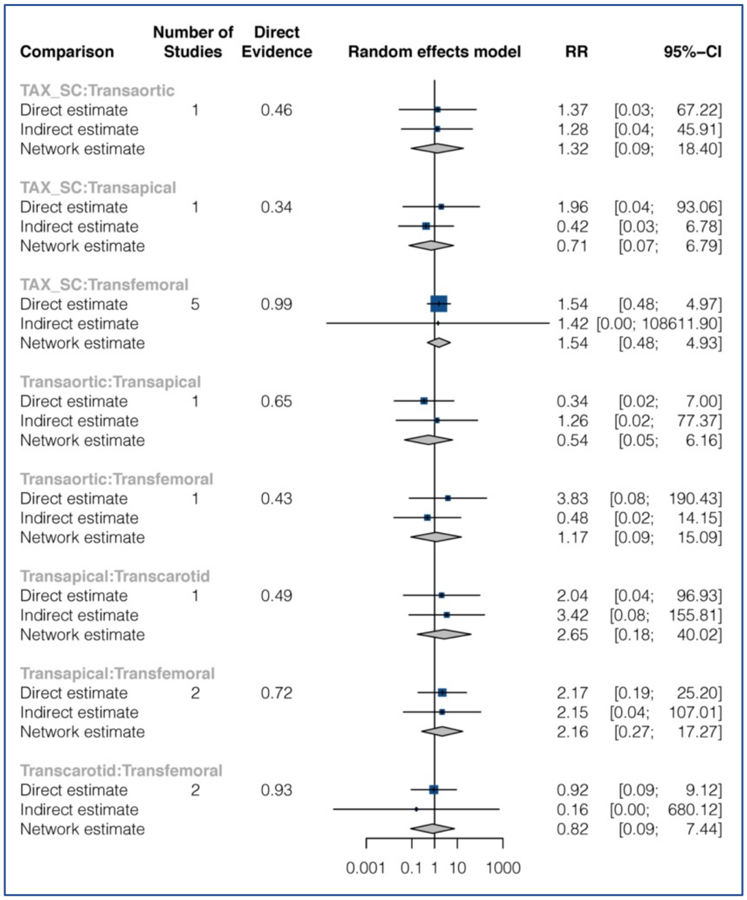

**h. Major Bleeding At 30-Days**

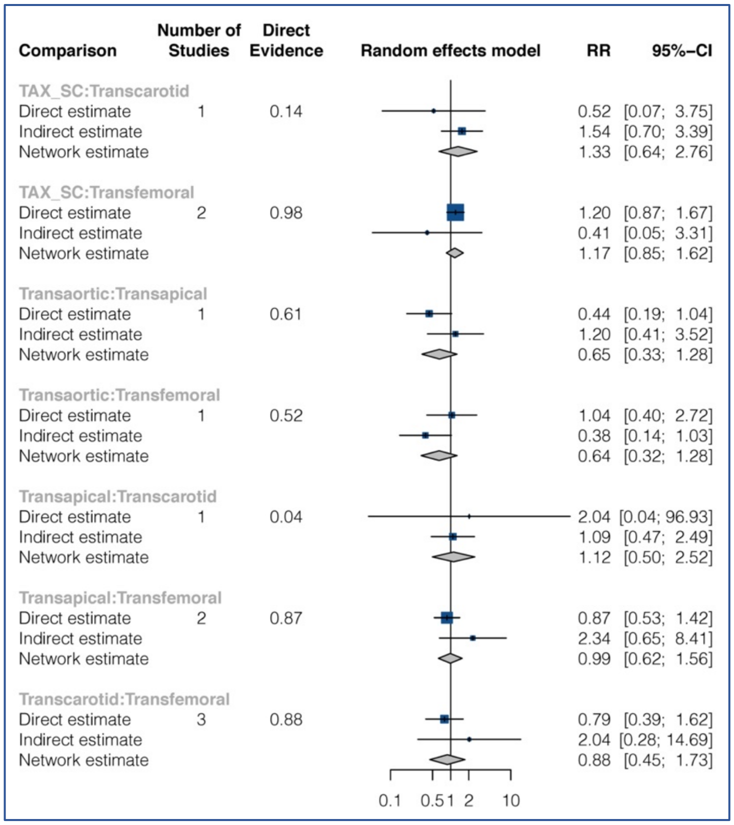


**i. MACCE At 30-Days**

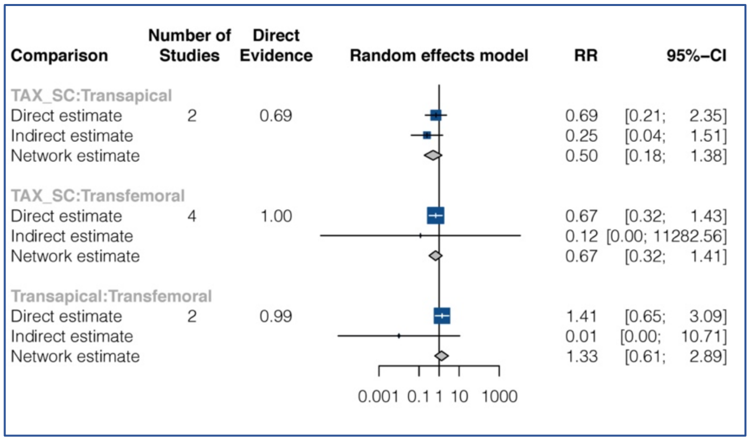

**eFigure-2**. Network graphs for the secondary outcomes.

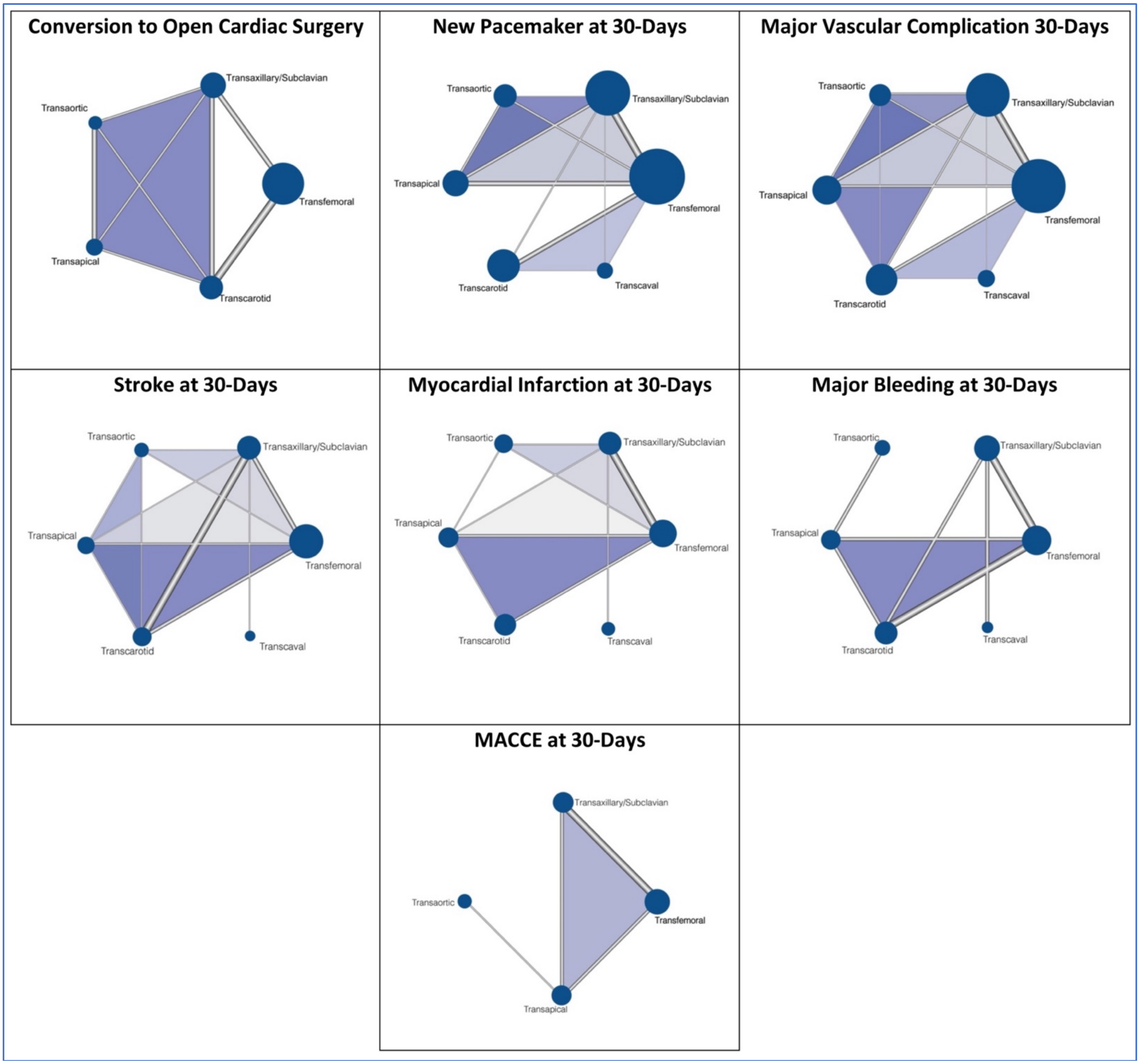


Each node (solid circle) represents an individual access site used in TAVR. The size of the node corresponds to the total number of patients included in each of the groups. The thickness of the edges (gray lines) connecting different nodes corresponds to the number of studies directly comparing a particular treatment pair.

**eFigure-3**. Rankograms for the secondary outcomes.

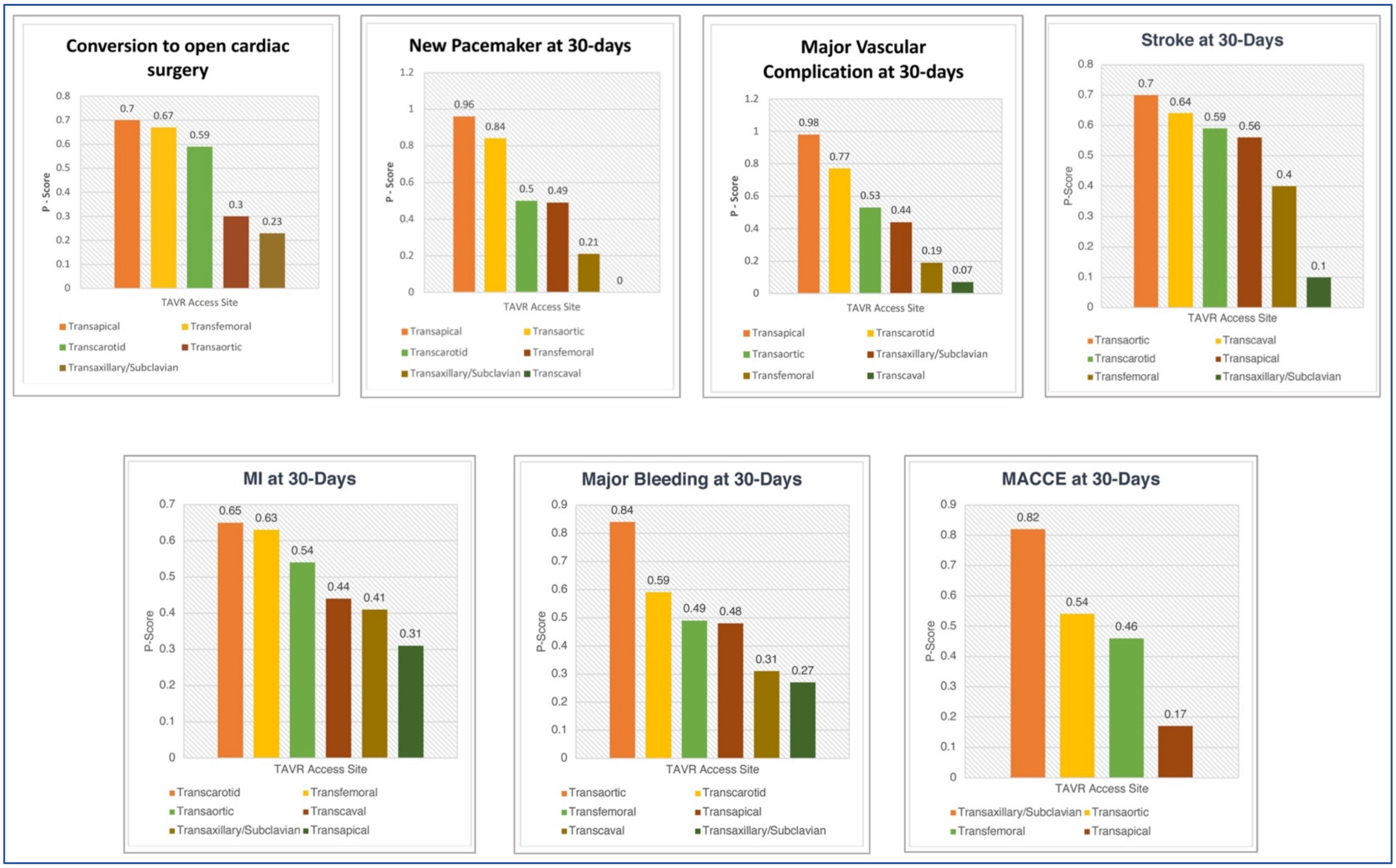
